# A Systematic Investigation of Detectors for Low Signal-to-Noise Ratio EMG Signals

**DOI:** 10.1101/2023.03.06.23286833

**Authors:** Monisha Yuvaraj, Priyanka Raja, Ann David, Etienne Burdet, Varadhan SKM, Sivakumar Balasubramanian

**Affiliations:** Department of Bioengineering, Christian Medical College Vellore, Tamil Nadu, India; Department of Applied Mechanics, Indian Institute of Technology Madras, Tamil Nadu, India; School of Electrical and Computer Engineering, Purdue University, Indiana, USA; Department of Mechanical Engineering, Indian Institute of Technology Madras, Tamil Nadu, India; Department of Bioengineering, Imperial College London, London, UK; School of Health and Rehabilitation Sciences, University of Queensland, Queensland, Australia

## Abstract

Active participation of stroke survivors during robot-assisted movement therapy is essential for sensorimotor recovery. Robot-assisted therapy contingent on movement intention is an effective way to encourage patients’ active engagement. For severely impaired stroke patients with no residual movements, a surface electromyogram (EMG) has been shown to be a viable option for detecting movement intention. Although numerous algorithms for EMG detection exist, the detector with the highest accuracy and lowest latency for low signal-to-noise ratio (SNR) remains unknown. This study, therefore, investigates the performance of thirteen existing EMG detection algorithms on simulated low SNR (0dB and -3dB) EMG signals generated using three different EMG signal models: gaussian, laplacian, and biophysical model. The detector performance was quantified using the false positive rate (FPR), false negative rate (FNR), and detection latency. Any detector that consistently showed FPR and FNR of no more than 20%, and latency of no more than 50ms, was considered an appropriate detector for use in robot-assisted therapy. The results indicate that the Modified Hodges detector – a simplified version of the threshold-based Hodges detector introduced in the current study – was the most consistent detector across the different signal models and SNRs. It consistently performed for ∼90% and ∼40% of the tested trials for 0dB and -3dB SNR, respectively. The two statistical detectors (Gaussian and Laplacian Approximate Generalized Likelihood Ratio) and the Fuzzy Entropy detectors have a slightly lower performance than Modified Hodges. Overall, the Modified Hodges, Gaussian and Laplacian Generalized Likelihood ratio, and the Fuzzy Entropy detectors were identified as the potential candidates that warrant further investigation with real surface EMG data since they had consistent detection performance on low SNR EMG data.

## Introduction

Substantial recovery of sensory-motor function after a stroke is possible with high-intensity and high-dosage movement training^1^. Rehabilitation robots can facilitate such high-intensity movement training while providing physical assistance to the user for completing movements consistently and precisely. About 30% of stroke survivors are severely impaired^2,3^ and require physical assistance to actively engage in movement training. While physical assistance from a robot can motivate such subjects to attempt and train movements, inappropriately timed or too much assistance ^4,5^ can lead to slacking, where patients reduce their voluntary effort and exploit robotic assistance to perform the movements. Inappropriately timed robotic assistance also alters the patient’s sense of agency or subjective awareness of control. This could lead to a lack of intrinsic motivation and attention, affecting motor learning and performance ^6^. Positive therapeutic effects have been observed only when patients actively engage in therapy^7– 10^. Jo et al.^11^ reported no improvement in clinical scales with passive range of motion therapy. Thus, active patient participation is an essential ingredient for sensorimotor recovery from movement therapy.

Robotic assistance contingent on a subject’s intention to move is an effective way to guide neuroplasticity^12^. However, how can the intention to move be detected in severely affected stroke patients with no residual movement? Electroencephalogram (EEG) based brain-computer interface (BCI) has been used to detect movement intention in order to trigger robotic movement assistance. The effectiveness of EEG-BCI in robot-assisted rehabilitation has been investigated in several studies^13–19^. Ramos et al. reported a 3 points improvement in the FMA score of the experimental group who were given robot-assisted therapy contingent on movement intention^17^. There are, however, several drawbacks to EEG-BCI systems:

1. They have a poor signal-to-noise ratio (SNR) with large trial-to-trial intra-subject variability^20^. The use of sophisticated signal processing techniques can increase the delay in intent detection, which can result in inappropriately timed robotic assistance leading to suboptimal recovery^1221^.
2. EEG-BCI modalities do not precisely identify which limb segment is intended to move due to its low information rate^21^.
3. Event-related desynchronization of the EEG sensorimotor rhythm may not necessarily reflect movement intention.^22^
4. Critical for practical use, current EEG-BCI systems are too complex and time-consuming for clinical work.

EMG could be a viable alternative to address these drawbacks of the EEG-BCI modality for robot-assisted therapy. EMG is a simple, robust, compact modality suitable for routine clinical use. In a recent study, we identified EMG as a potential alternative to EEG-BCI to detect movement intention from severely affected stroke patients without residual movement^23^. About 70% (22 out of the 30) of the study participants had residual EMG in the forearm muscles that showed a consistent increase in amplitude with wrist/finger movement attempts.

However, the study reported poor agreement between the EEG and EMG modalities for detecting movement intention. The authors suggested that this discrepancy could be because of the simple root mean square detector with temporal thresholding that was used. This detector may not optimally pick up low SNR surface EMG signals^23^ expected from severe patients with no residual movements.

Numerous EMG detection algorithms have been proposed for the automatic detection of EMG onset^24–31^. The review article by Staude et al. ^24^ compared different EMG detector types to identify the best detectors for identifying EMG onset ^24^. However, this work was done on high SNR (3dB and 6dB) simulated EMG signals generated by bandpass-filtering white Gaussian noise, and it only investigated detectors reported till 2001. Other detectors have been reported in the last 20 years, and a systematic characterization of existing EMG detector types on low SNR EMG signals (generated using different signal models) is still lacking. Identifying an optimal detector is essential for further exploring the use of EMG-based movement intent detection for robot-assisted therapy in severely affected stroke subjects with no residual movements.

Our goal in this study is to systematically compare the detection accuracy and latency of existing EMG detectors on low SNR EMG signals to identify the most promising detectors, while eliminating the ones with poor performance, for further investigation. To this end, we will here:

- Generate simulated low SNR (0dB and -3dB) EMG data using two phenomenological (Gaussian and Laplacian) models and a biophysical model to evaluate the performance of the different detector types.
- Define an appropriate cost function considering the detection accuracy and latency to evaluate the performance of the different detector types.
- Compare the performance of the different detector types on simulated surface EMG signals from the three signal models for two different SNRs (0dB and - 3dB) and identify the most appropriate detector type(s).

We note that the detectors studied in this work produce a binary output suitable for on-off control of robotic assistance during therapy. This is in contrast to previous studies exploring continuous control paradigms using EMG amplitude estimates on less severely affected stroke patients with some residual limb movements^32,33^ However, for severely affected patients with no residual movements, who may or may not have detectable EMG, an on-off control scheme might be the easiest modality for human-robot interaction. As patients gain better control over their muscles, continuous control schemes might be appropriate as patients gain better control of their muscles that are still too weak to produce residual movements.

## Methods

Neurorehabilitation training consists of repeating specific movements of interest punctuated by periods of rest. A typical training session will involve several “trials” of a particular movement, each with a period of rest (rest-phase) followed by a period of movement/movement attempt (move-phase). In EMG-triggered robot-assisted therapy, robotic assistance is triggered whenever EMG is detected in real-time in the target muscle during the move phase of a trial. In this paper, EMG always refers to surface-recorded muscle activity, as this modality will most likely be employed in routine robot-assisted therapy. The rest of this section starts with the formal definition of the signal processing problem solved by an EMG detector, followed by the details of the simulated EMG signals, a description of the general structure of EMG detectors, and the approach used to compare the performance of the different detectors.

### The formal definition of the signal processing problem

Let *x*_*i*_[*n*], 0 ≤ *n* < *N*_*t*_ be the recorded signal from a target muscle during the *i*^*th*^ trial; *n* is the sampling instant, where *n* = 0 is the start of a trial, *N*_*t*_ is the number of data points from each trial. Let *N*_*r*_ and *N*_*m*_ be the number of samples in the rest- and move-phases of a trial, respectively, then *N*_*t*_ = *N*_*r*_ + *N*_*m*_. The time segments 0 ≤ *n* < *N*_*r*_ and *N*_*r*_ ≤ *n* < *N*_*t*_ correspond to the rest-phase and move-phase of the trial, respectively.

### Problem definition

To detect the presence of EMG in realtime in the move-phase of a trial using only the current and past EMG data {*x*[*K*]}, 0 ≤ *K* ≤ *n* from the start of the trial. Let *y*[*n*] represent the binary output of the EMG detector at the current sampling instant *n*,

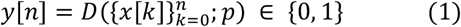

where, *D*(·) is the detector function that maps the EMG signal 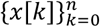, from the start of a trial to a binary output corresponding to the presence or absence of EMG at the current time instant *n*; *p* is the set of detector parameters that control the detector’s performance. The function *D*(·) is often a complex mathematical operation consisting of a series of simpler operations performed on the EMG data to produce the binary output. This binary output can be used as a simple on/off control of robotic assistance by severely affected patients to relearn movement initiation^34,35^.

### Simulation of Surface EMG Signal

The analysis of the different EMG detectors was performed using simulated EMG data. To generate this simulated EMG, we assume that:

- the measurement noise has a fixed variance throughout the experiment.
- the muscle is fully relaxed in the rest-phase of any trial, i.e., there is no EMG activity from the target muscle during the rest-phase, and
- the muscle is activated at a constant level for the entire duration of the move-phase.

These assumptions were made to evaluate the EMG detectors under the conditions that: (a) The EMG signal has a fixed signal-to-noise ratio (SNR) in the move-phase, and (b) All other intra- and inter-trial variabilities in the EMG signal characteristics is minimized. The detectors that perform poorly under these ideal conditions will likely perform worse with real EMG data from patients.

We simulated 100 trials of EMG data with an individual trial duration of 13 seconds (8s and 5s for rest- and move-phases, respectively). The sampling frequency of the simulated signal was set to 1000Hz. Three different EMG signal generation models were employed in the current analysis – two phenomenological (Gaussian and Laplacian) models and one biophysical model. The phenomenological models were based on the work of De Luca^36^ where a surface EMG signal from a muscle activated at a fixed level can be treated as zero-mean white noise followed by a shaping filter (electrode properties); this model is widely accepted in the literature^24,37–40^. The exact probability density of the EMG signal depends on the muscle activation level, with high levels of activation following a Gaussian distribution^37,38^. However, at low levels of muscle activation, EMG signals have been reported to follow a distribution that lies between a Gaussian and a Laplacian distribution^41^. Therefore, to ensure that the detectors are tested with the appropriate signals, we generated data using both white Gaussian and white Laplacian signals, resulting in two phenomenological models.

### Phenomenological Models: Gaussian and Laplacian

The first step in this model is the generation of the zero-mean unit-variance white Gaussian and Laplacian noise *e*[*n*]. A step change in the signal variance, within a trial, at the transition between the rest- and move-phases of a trial was obtained by multiplying *e*[*n*] by *σ*[*n*]:

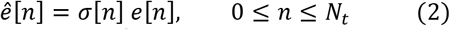

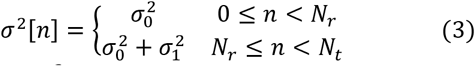

where 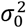 and 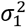 are the noise and signal variances, respectively. The noise variance is always set to 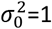 in this analysis, and the signal variance is chosen based on the desired signal-to-noise ratio (SNR) in the move-phase. The signal 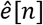 is then zero-phase bandpass filtered (8^th^ order FIR bandpass filter with cut-off frequency 10 Hz and 450 Hz) to have a signal with spectral characteristics like a surface EMG signal.

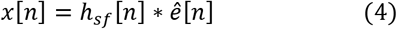

where, *h*_*sf*_[*n*] is the impulse response of the bandpass or shaping filter, and *x*[*n*] is the generated EMG signal that is used for the analysis. The signal-to-noise ratio (SNR) of this simulated EMG signal in the move-phase is given by,

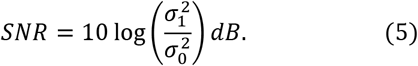

### Biophysical Model

In addition to the phenomenological models, we also wanted to test the detectors on more realistic data based on the biophysics of the EMG signal, accounting for the physiological origin of the electrical muscle activity and the recording electrode geometry. In this paper, the biophysical model proposed in Devasahayam.S^42^ was employed to generate the simulated EMG data. Assuming a linear, isotropic volume conduction model, a simple muscle geometry with parallel muscles fibres ignoring the effects due to the finite muscle fibre length, the EMG recorded by a bipolar electrode configuration can be approximated using the following expression,

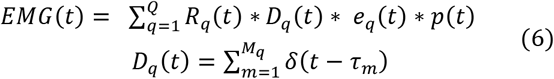

where *Q* is the number of motor units in the muscle, *R*_*q*_(*t*) is the impulse train signal arriving at the *q*^*th*^ motor unit through its corresponding motor neuron, *M*_*q*_ is the number of muscle fibers in the *q*^*th*^ motor unit, *e*_*q*_(*t*) is the approximate electrode transfer function between the *q*^*th*^ motor unit and the recording electrodes, and *p*(*t*) is the single fibre action potential, which is assumed to be the same for all fibres. The full details of the model can be found in Devasahayam.^42^ with the associated parameters provided in Table S3 of the supplementary material.

The EMG simulator developed by Devasahayam. was employed in the current work to generate the simulated EMG signals^42^. A bipolar surface electrode configuration with a 10mm interelectrode distance was considered. The simulator takes in the muscle force level as its input and computes the corresponding firing pattern for the motor units. In the current study, the force levels from the muscle were set to 0N in the rest-phase (no muscle activation) and 10N in the move-phase (average firing rate of 16.4 Hz for the muscle). The simulator generated pure muscle activity *x*_*pure*_[*n*] recorded by the chosen electrode configuration. The force level for the muscle in the move-phase was chosen empirically to ensure that the temporal profile of the simulated EMG signal (*x*_*pure*_[*n*], 0 ≤ *n* ≤ *N*_*t*_) visually resembled that of real surface EMG signals. A zero-mean white Gaussian noise *e*[*n*] of fixed noise variance 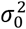 was added to *x*_*pure*_[*n*] to introduce measurement noise. The noise variance was chosen based on the signal power 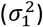 of *x*_*pure*_[*n*] in the move-phase to obtain a signal with the desired SNR:

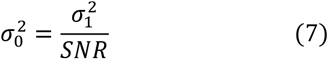

Following this, the noisy signal (*x*_*pure*_[*n*] + *e*[*n*]) is bandpass filtered (8th order non-causal FIR filter) between 10 Hz and 450 Hz cut-off frequencies:

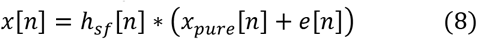

where *h*_*sf*_ [*n*] is the impulse response of the bandpass shaping filter. The characteristics of EMG generated from these three models are shown in Fig.1. The current study employed two different SNRs of 0dB and -3dB.

**Figure 1:**
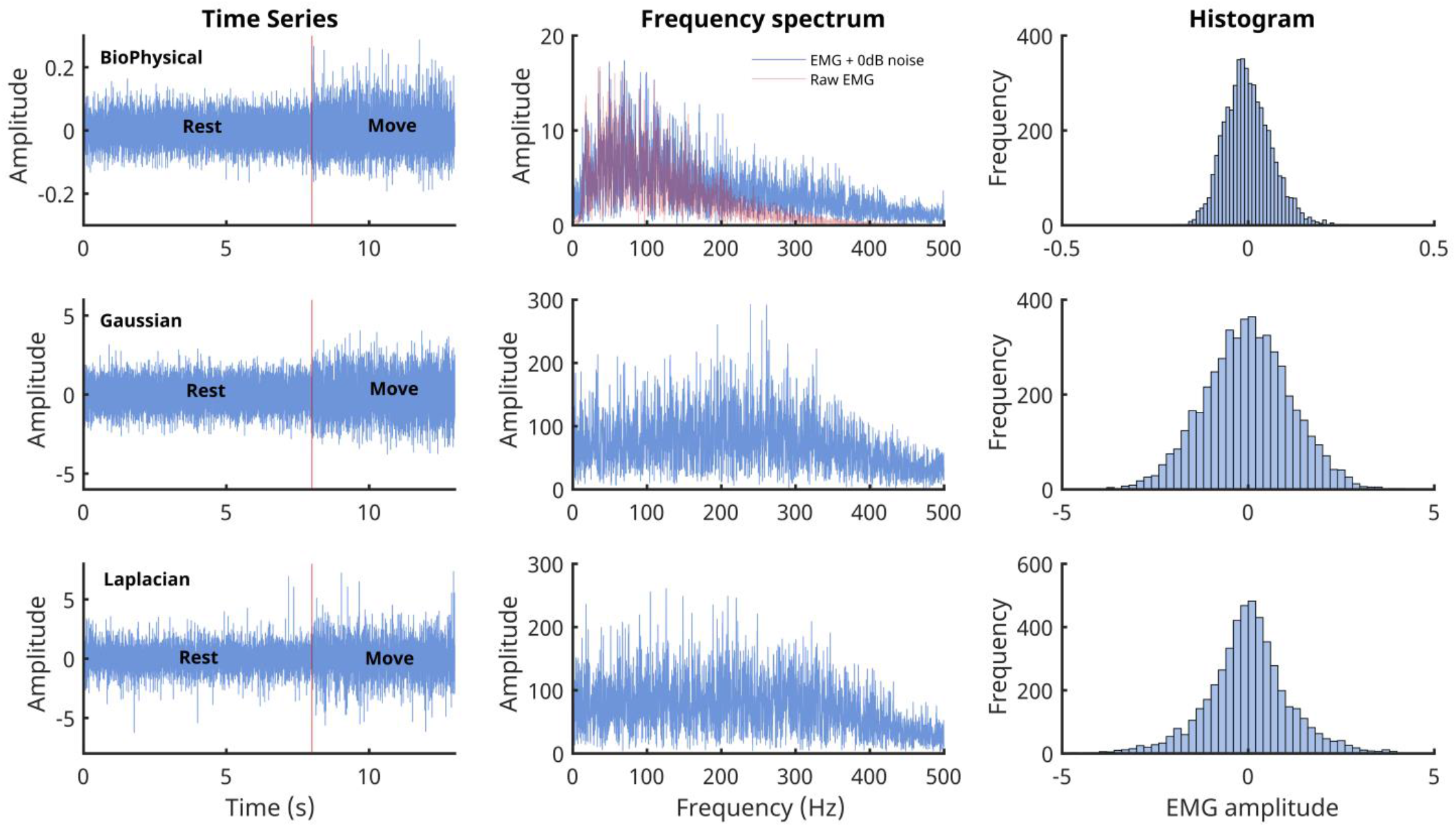
Characteristics of the EMG signals generated from the three models. The three rows correspond to the three different signal models: B Biophysical in the top row, Gaussian in the middle, and Laplacian in the bottom row. The left most column shows the time series of the simulated 13 seconds of data with the first 8 seconds corresponding to the rest phase and the rest 5 seconds to the move phase. The middle column shows the corresponding Fourier magnitude spectrum of the 5 seconds of move phase data. The right column displays the estimate of the probability density functions of the 5 seconds of move phase data from the three models.

### Detection Algorithms

The general structure for EMG detectors proposed by Staude et al.^24^ is shown in Fig.2, which consists of three steps carried out sequentially to map the given real-time EMG data into a binary output:

**Figure 2:**
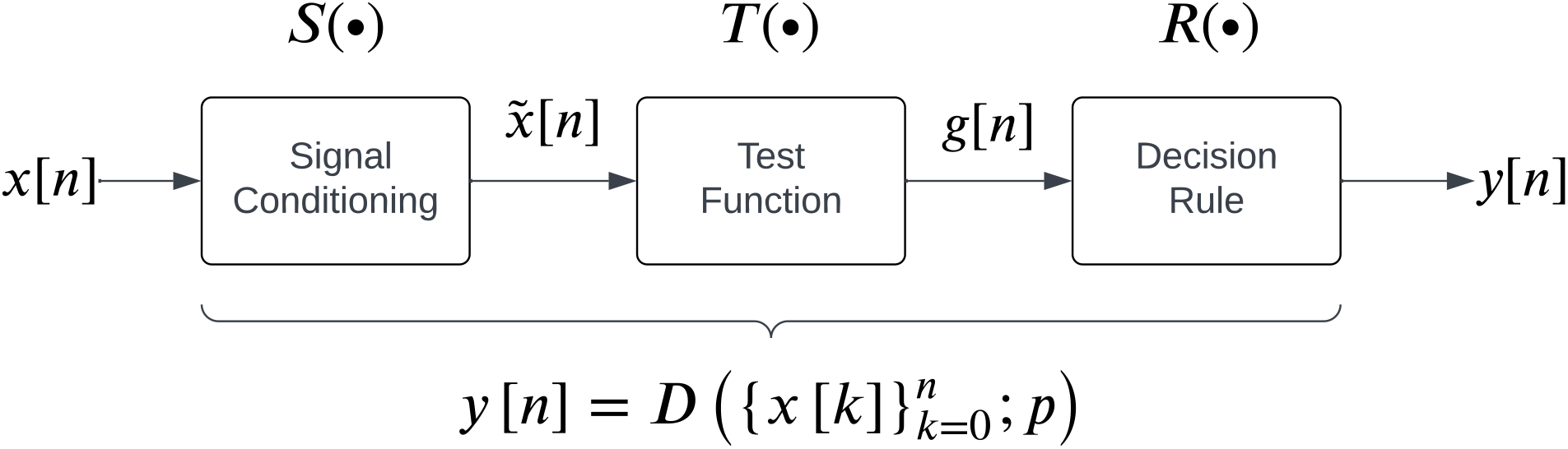
A general structure for EMG detectors as proposed by Staude et. al.^24^

1. **Signal conditioning** is the first step to improve EMG signal quality for better detection, often involving high-pass filtering for movement artefact removal. Some detectors might employ additional filtering operations, such as adaptive whitening for stable EMG amplitude estimation^43,44^. The conditioned signal is represented by

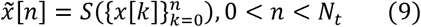

where *S*(·) represents the mathematical operation performed by the signal conditioning step.
2. **Test function computation transforms** 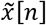 into a scalar variable or feature that can distinguish the presence or absence of EMG. The test function *g*[*n*] is computed at the current time instant *n* over a causal window of size *W*:

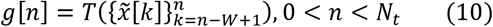

Some examples of test functions in the literature include the moving average of 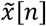, *χ*^2^ test variable, likelihood ratio etc.
3. A **decision rule** is applied on the test function *g*[*n*] by comparing it to a threshold *h* to identify the presence/absence of an EMG signal:

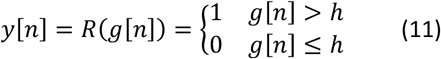

Some detectors employ a more sophisticated decision rule, such as double thresholding, to control the false positive rates of detection^45,46^.

We note that each detector has a set of parameters associated with it. The current study compares the performance of 13 detector types reported in the literature which can be implemented in real-time, listed in Table 1. Each detector type fits into the general structure shown in Fig. 2. The different parameters associated with these detector types are also provided in Table 2. A detailed description of the individual detector types and the algorithms for their implementation are provided in the supplementary material (Table S1).

**Table 1:**
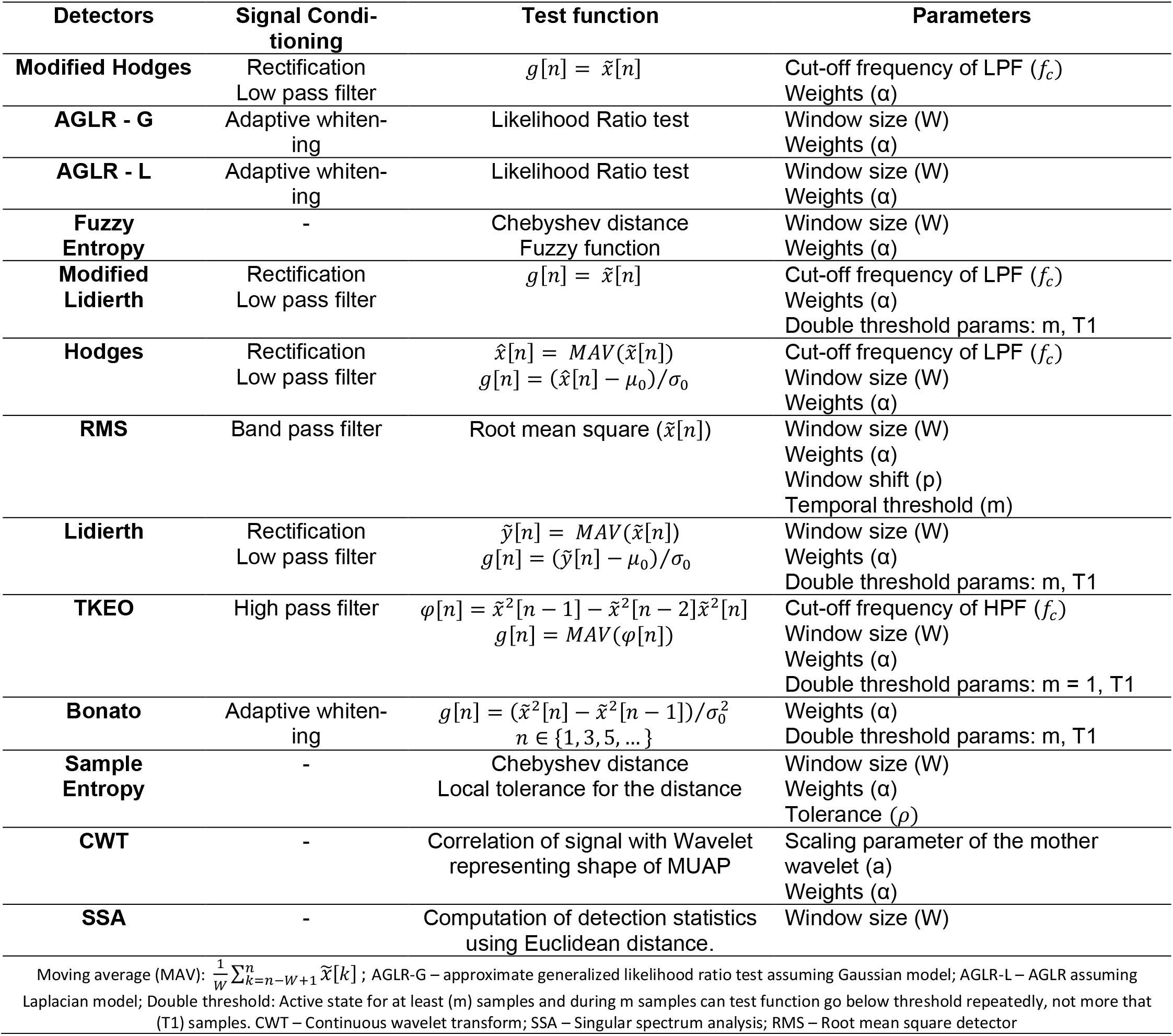
Description of the structure of the 13 detectors investigated in the current study, along with the different parameters associated with the individual detectors.

**Table 2:** Optimal parameters for the different detector types for the different SNRs and signal model. These parameters were identified using procedure described in Algorithm 1 on the 50 trials from the training datasets.

| Detector Type | Parameters | SNR 0dB |  |  | SNR -3dB |  |  |
| --- | --- | --- | --- | --- | --- | --- | --- |
|  |  | Gaussian | Laplacian | Biophysical | Gaussian | Laplacian | Biophysical |
| **Modified Hodges** | Weight | 1 | 1 | 1 | 1 | 1 | 1 |
|  | LPF cut-off (Hz) | 7.5 | 7.5 | 4.5 | 6.5 | 5.5 | 4.5 |
| **AGLR-G** | Window Size (s) | 0.1 | 0.1 | 0.15 | 0.1 | 0.15 | 0.15 |
|  | Weight | 2 | 1 | 3 | 1 | 1 | 1 |
| **AGLR-L** | Window Size (s) | 0.1 | 0.1 | 0.1 | 0.1 | 0.15 | 0.2 |
|  | Weight | 2 | 1 | 1 | 1 | 1 | 1 |
| **Fuzzy Entropy** | Window Size (ms) | 60 | 80 | 100 | 90 | 70 | 100 |
|  | Weight | 1 | 1 | 2 | 1 | 1 | 1 |
| **Modified Lidieth** | LPF cut-off | 9.5 | 9.5 | 9.5 | 9.5 | 9.5 | 7.5 |
|  | Weight | 1 | 1 | 1 | 1 | 1 | 1 |
|  | m | 20 | 25 | 5 | 25 | 55 | 25 |
|  | T1 | 30 | 30 | 30 | 30 | 60 | 30 |
| **Hodges** | Window Size (s) | 0.1 | 0.1 | 0.1 | 0.1 | 0.1 | 0.1 |
|  | Weight | 1 | 1 | 1 | 1 | 1 | 1 |
|  | LPF cut-off (Hz) | 9.5 | 9.5 | 9.5 | 9.5 | 8.5 | 9.5 |
| **RMS** | Window Size (s) | 0.12 | 0.12 | 0.12 | 0.12 | 0.12 | 0.12 |
|  | Weight | 1 | 1 | 1 | 1 | 1 | 1 |
|  | Window shift (ms) | 40 | 40 | 40 | 40 | 40 | 40 |
|  | Time threshold (ms) | 40 | 40 | 40 | 40 | 40 | 40 |
| **Lidieth** | Window Size (s) | 0.1 | 0.1 | 0.1 | 0.1 | 0.1 | 0.1 |
|  | Weight | 1 | 1 | 1 | 1 | 1 | 1 |
|  | m | 10 | 10 | 5 | 20 | 25 | 25 |
|  | T1 | 30 | 30 | 30 | 30 | 30 | 30 |
| **TKEO** | HPF cut-off (Hz) | 5 | 5 | 5 | 15 | 20 | 15 |
|  | Window Size (s) | 0.1 | 0.1 | 0.1 | 0.1 | 0.1 | 0.1 |
|  | Weight | 1 | 1 | 1 | 1 | 1 | 1 |
|  | T1 | 30 | 30 | 30 | 30 | 30 | 30 |
| **Bonato** | Weight | 1 | 2 | 2 | 2 | 1 | 2 |
|  | m | 10 | 25 | 20 | 20 | 20 | 25 |
|  | T1 | 30 | 30 | 30 | 30 | 60 | 30 |
| **Sample Entropy** | Window Size (ms) | 50 | 50 | 50 | 50 | 50 | 50 |
|  | Weight | 1 | 1 | 1 | 1 | 1 | 1 |
|  | Tolerance for distance | 0.5 | 1.5 | 0.5 | 0.5 | 1.5 | 0.5 |
| **CWT** | Weight | 1.1 | 1.2 | 1 | 1.1 | 1 | 1.4 |
| **SSA** | Window Size (ms) | 52 | 50 | 50 | 50 | 50 | 50 |

### A Measure of Detector Performance

The simulated EMG data from the three different signal models and the two different SNRs were used to evaluate the performance of the different detector types. Each trial of EMG signal (13 seconds long) was input to the different detectors to compute binary output indicating the presence or absence of EMG signal. The performance of a detector must consider the accuracy (false positive and false negative rate) and the detection latency. These are computed from the output *y*[*n*] of each trial (Fig. 3), where the EMG signal from each trial was analyzed in the following three steps:

**Figure 3:**
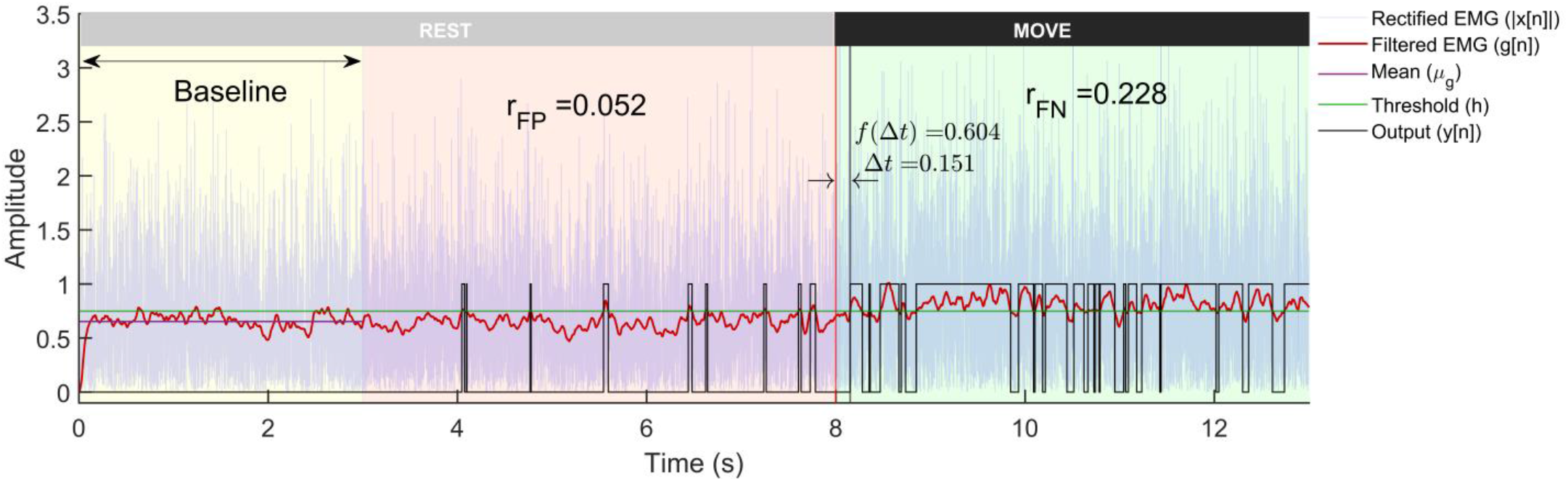
A representative example of a trial from the Gaussian signal model with -3dB SNR run through the Modified Hodges detector. The plot shows the rectified EMG signals, its lowpass filtered output, and the binary output from the detector. The trial is 13 seconds long with first 8 seconds corresponding to the rest-phase and the rest 5 seconds to the move-phase. The rest-phase is further divided into the baseline phase (yellow background) that is used computing the threshold *h*, and the remaining rest-phase (red background) is used for computing *r*_*FP*_. The move-phase (green background) is used to compute Δ*t* and *r*_*FN*_.

1. the first three seconds (0–3s) of the rest-phase data is used for estimating the threshold *h* for detection:

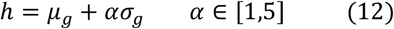

where *μ*_*g*_ and *σ*_*g*_ are the mean and standard deviation of the test function in this period.
2. the remaining 5 seconds of the rest-phase are used to estimate the false positive rate (*r*_*FP*_).
3. the 5 seconds of the move-phase are used to estimate the false negative rate (*r*_*FN*_) and the detection latency (Δ*t*).

We defined a performance measure to compute a single number referred to as the *cost of detection* that considers the false positive rate *r*_*FP*_, the false negative rate *r*_*FN*_, and the detection latency (Δ*t*). Let ***c*** ≜ [*r*_*FP*_ *r*_*FN*_ *f*(Δ*t*)]^*T*^ be the cost vector associated with the output *y*[*n*] of the detector for a particular trial. We define the *cost* of detection *C* as the infinity norm of the cost vector ***c***.

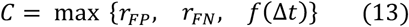

providing the worst-case performance of the detector on the given trial.

The false positive rate *r*_*FP*_ is defined as the ratio of the number of 1s in the detector output *y*[*n*] in the rest-phase of a trial, and the false negative rate *r*_*FN*_ is defined as the ratio of the number of 0s in *y*[*n*] in the move-phase.

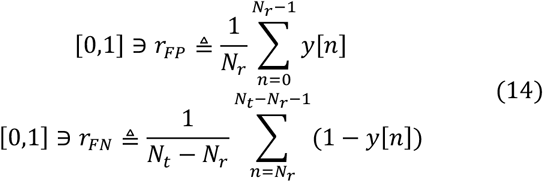

From Fig.3, the detection latency is defined as the time delay from the start of the move-phase to when the detector output goes to 1:

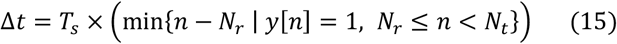

where *T*_*s*_ is the sampling period of data in milliseconds, and Δ*t* ∈ [0, 5000]*ms*. The cost due to this latency is quantified by the function *f*(Δ*t*) that maps Δ*t* to a real number in the closed interval between 0 and 1:

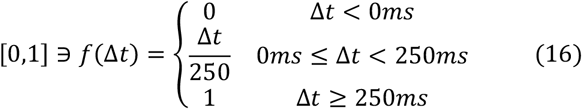

Latencies between 0 to 250ms have linearly increasing costs while the ones above 250ms are considered as bad as 250ms. Based on the definitions of *r*_*FP*_, *r*_*FN*_, and *f*(Δ*t*), *C* ∈ [0,1]. A detector with a consistently lower cost of detection *C* would be considered a better detector.

### Comparing Different Detector Types

A detector’s performance or cost is determined by the SNR of the input signal, the detector type, and its associated parameters. Thus, for a fixed SNR input signal, comparing two detector types must be done only after controlling for the influence of their corresponding detector parameters. In the current work, this was done by first choosing the optimal parameters for each detector type, before comparing different detector types. The optimal parameters for a detector type were selected by first splitting the 100 movement trials of simulated EMG data into training and validation datasets with 50 trials each. This was done for both SNRs (0dB and -3dB) and for all three signal models (gaussian, laplacian, and biophysical). The training dataset was used to identify the optimal parameter values for the different detector types, i.e. the values of the parameter combination that consistently resulted in the least cost for the detector on the training dataset. The exact procedure is given in Algorithm 1, while details are provided in the supplementary materials.

After identifying the optimal parameter combination for each detector type, the optimal parameter values were used to run the detector on the 50 trials of the validation dataset, which resulted in the validation cost set 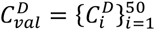 for the detector type *D*. The cost set from the different detector types were compared using two-way ANOVA with the detector type and signal SNR as the two factors for each of the three signal model. The complete code for the analysis can be found <u>here</u>.

## Results

The entire analysis reported in this paper was implemented in MATLAB R2020. A sample of the individual trials from the three EMG signal models is depicted in Fig. 1. A sample output of the different processing stages of the Modified Hodges detector from a Gaussian EMG signal trial is shown in Fig. 3. The Modified Hodges detector filters (2nd order Butterworth low pass filter) the rectified raw EMG signal (blue colored trace in Fig.3). This lowpass filtered signal is the test function of the detector (red-colored trace in Fig. 3). The threshold *h* for this trial is shown by the green-colored horizontal line in Fig. 3. The output of the detector (black colored trace in Fig.3.) is 1 whenever the test function crosses the threshold, it is 0 otherwise. The figure also shows the values of *r*_*FP*_, *r*_*FN*_, Δ*t*, and *f*(Δ*t*) for the trial.

### Optimal parameters for the different detector types

The thirteen detector types were compared after choosing the optimal parameter set for each detector type using the training dataset of 50 trials for each of the six combinations of the three signal models and two SNRs. This procedure is depicted in Fig. 4 for the Modified Hodges detector for the 0 dB SNR Gaussian signal model, which shows the outcomes from the different steps in the optimization process described in Algorithm 1. Fig. 4(a) shows the histograms of the cost *C* for the different parameter combinations, in light blue traces. These histograms are estimated from the cost values 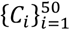 obtained from the 50 trials in the training dataset for the different combinations of the detector parameters. The scatter plot of the median *c*_*med*_ and inter-quartile range *c*_*iqr*_ of these histograms are shown in Fig. 4(b). The choice of the best parameter for the detector was determined to be the one with the least euclidian norm 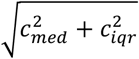 which is shown as the red circle in Fig. 4(b); its corresponding histogram is shown in the thick red trace in Fig. 4(a). Fig. 4(c) shows the marginal histograms of the individual contributors (*r*_*FP*_, *r*_*FP*_, *f*(Δ*t*)) to the cost *C* for the optimal parameter combination for the Modified Hodges detector. The values of the optimal parameters for the different detector types are listed in Table 2.

**Figure 4:**
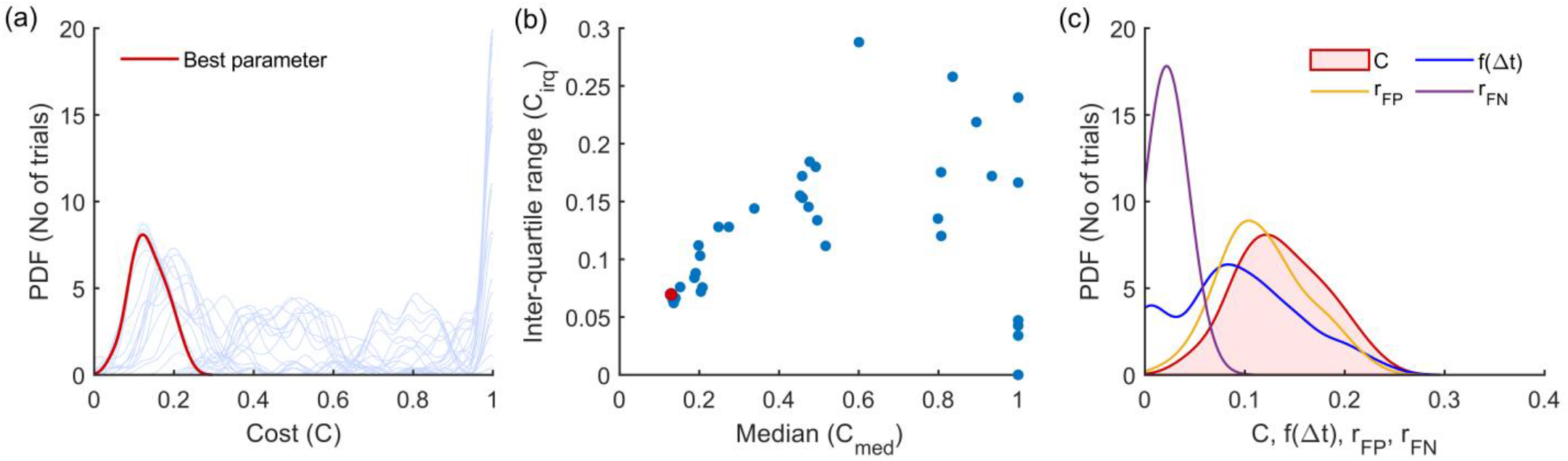
Outcomes of the parameter optimization process for the Modified Hodges detector as described in Algorithm 1. (a) Estimated probability density function of cost for the different combinations of parameters (in light blue traces). The red trace corresponds to the cost of the optimum parameter combination. (b) Scatter plot of median vs IQR for the cost of different parameter combinations. (c) Estimated histograms of latency, false positive rate, false negative rate and cost of the optimum parameter.

### How do the different detector types perform on the different signal models and SNRs?

The performance of the different “optimal” detector types, i.e., detectors using the optimal parameter values, were compared using the 50 trials from the validation datasets. The boxplot of the performance of these different detector types for the three different signal models – gaussian, laplacian, and biophysical – are shown in Fig. 5(a), (b), (c), respectively; each of these subplots displays the performance for the 0dB and -3dB SNRs in red and blue boxplots, respectively. Note that the order of the depiction of the different detectors is in terms of the increasing average cost across the three signal models and two SNRs; the detectors on the left are better than the ones on the right in an average sense. A two-way ANOVA on the effect of the detector type and SNR on performance revealed a significant difference between the detector types (p < 0.001) and SNRs (p < 0.001) for all three signal models. The test revealed a significant interaction between the factors for all three models (biophysical: p < 0.0001; Gaussian: p < 0.0001; Laplacian: p < 0.0001). These statistical results confirm the results shown in the boxplots in Fig. 5, where the performance is different among the detector types, with consistently poorer performance for -3dB compared to 0dB. The costs for both 0dB and 3dB appear to be lower for the biophysical model compared to the Gaussian and Laplacian models.

**Figure 5:**
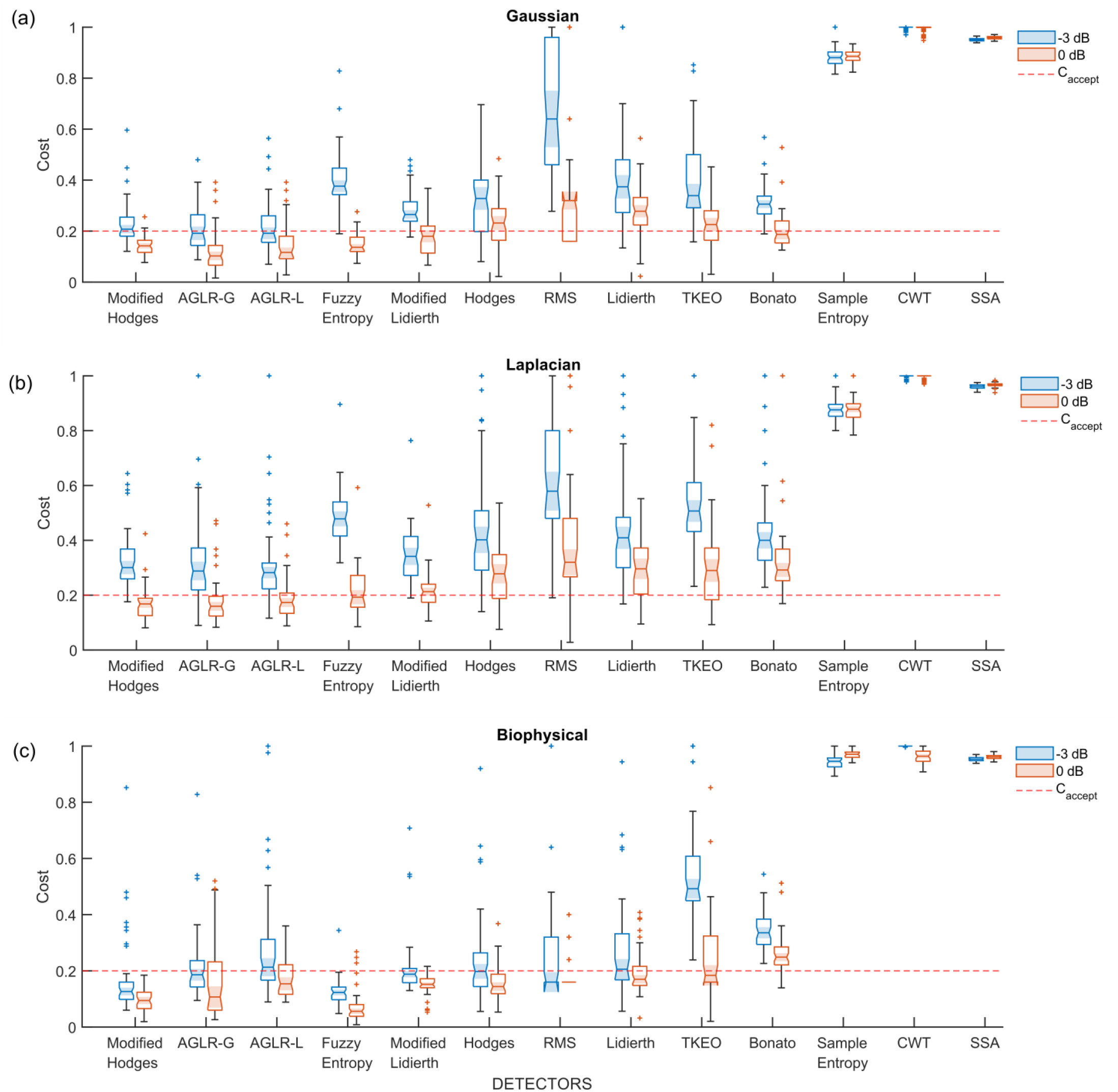
Boxplot of cost of performance of the different detectors in the validation datasets from the different signal models and SNRs. All detectors shown in this plot use the optimal detector parameter sets optimized on the training dataset. (a) Cost of detection for the Gaussian signal model, (b) Laplacian signal model and (c) Biophysical signal model. The red and blue colored boxplots are of 0dB and -3 dB SNR, respectively. The red dashed line is the acceptable cost *C*_*accept*_ = 0.2.

Most detector types perform similarly except for the Sample Entropy, CWT, and SSA detector types which perform worse across the different signal models and SNRs. Among the other detector types – the Modified Hodges, the AGLR-G, and AGLR-L detectors – have almost similar costs for the different signal models and SNRs. The other detector types – RMS, Hodges, Bonato, Lidierth, Modified Lideirth, TKEO, and Fuzzy Entropy – have slightly higher costs for one or more specific signal models and SNRs. We note that the Fuzzy Entropy detector performs very well on the biophysical signal model for both SNRs.

### Which detector types have an acceptable cost?

The choice of appropriate detector type(s) for use in robot-assisted therapy requires the specification of an acceptable cost of detection *C*_*accept*_. To this end, we specify the upper limits for the false positive rate, false negative rate, and the latency of detection as *C*_*accept*_ = 0.2, which corresponds to a detector with the following cost components:

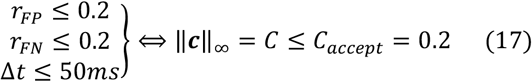

Any detector type with costs consistently lower than *C*_*accept*_ would be deemed an appropriate detector for use in robot-assisted therapy. To determine the detector types with consistently lower costs than *C*_*accept*_, we computed the proportion 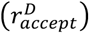 of the 50 validation trials with acceptable *C* ≤ *C*_*accept*_ for the detector *D* for the three signal models and two SNRs using

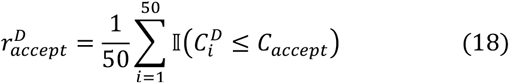

where 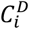 is the cost of detection for the *i*^*th*^ validation trial for detector *D*. The value of 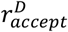 for the different detector types is shown in Table 3, where the cells with 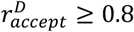 are highlighted. We can observe there that:

**Table 3:**
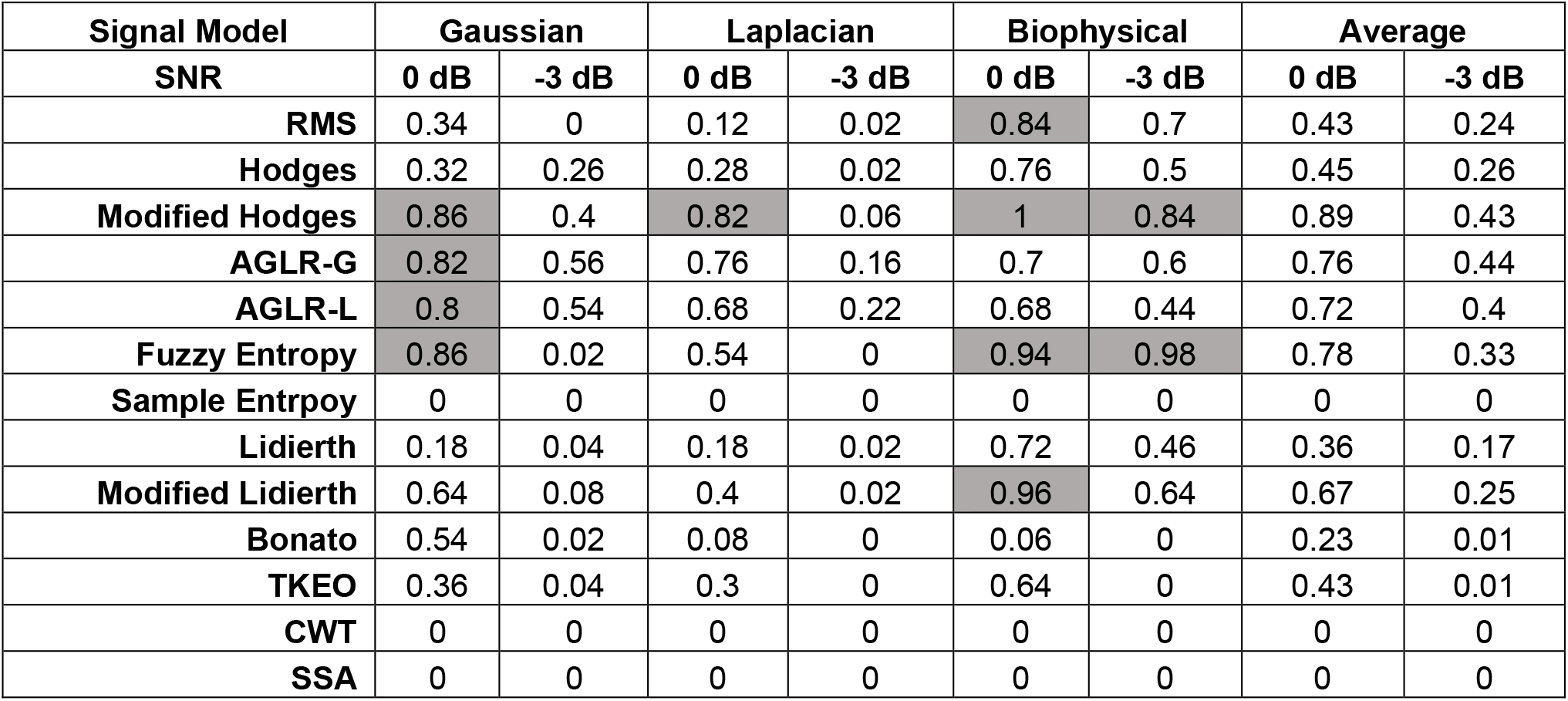
The proportion of the 50 validation trials with cost less than the acceptable cost of 0.2 for different detector types, signal models, and SNRs. The cells with proportions greater or equal to 0.8 are highlighted in gray.

| Signal Model | Gaussian |  | Laplacian |  | Biophysical |  | Average |  |
| --- | --- | --- | --- | --- | --- | --- | --- | --- |
| SNR | 0 dB | -3 dB | 0 dB | -3 dB | 0 dB | -3 dB | 0 dB | -3 dB |
| **RMS** | 0.34 | 0 | 0.12 | 0.02 | 0.84 | 0.7 | 0.43 | 0.24 |
| **Hodges** | 0.32 | 0.26 | 0.28 | 0.02 | 0.76 | 0.5 | 0.45 | 0.26 |
| **Modified Hodges** | 0.86 | 0.4 | 0.82 | 0.06 | 1 | 0.84 | 0.89 | 0.43 |
| **AGLR-G** | 0.82 | 0.56 | 0.76 | 0.16 | 0.7 | 0.6 | 0.76 | 0.44 |
| **AGLR-L** | 0.8 | 0.54 | 0.68 | 0.22 | 0.68 | 0.44 | 0.72 | 0.4 |
| **Fuzzy Entropy** | 0.86 | 0.02 | 0.54 | 0 | 0.94 | 0.98 | 0.78 | 0.33 |
| **Sample Entropy** | 0 | 0 | 0 | 0 | 0 | 0 | 0 | 0 |
| **Lidierth** | 0.18 | 0.04 | 0.18 | 0.02 | 0.72 | 0.46 | 0.36 | 0.17 |
| **Modified Lidierth** | 0.64 | 0.08 | 0.4 | 0.02 | 0.96 | 0.64 | 0.67 | 0.25 |
| **Bonato** | 0.54 | 0.02 | 0.08 | 0 | 0.06 | 0 | 0.23 | 0.01 |
| **TKEO** | 0.36 | 0.04 | 0.3 | 0 | 0.64 | 0 | 0.43 | 0.01 |
| **CWT** | 0 | 0 | 0 | 0 | 0 | 0 | 0 | 0 |
| **SSA** | 0 | 0 | 0 | 0 | 0 | 0 | 0 | 0 |

- All detectors perform poorly for the -3dB Laplacian signal model. The highest value of 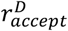 is 0.22 for this signal model, which interestingly is from the AGLR-L detector designed for the Laplacian signal. Many detectors perform a little better with higher 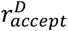 values for the -3dB Gaussian and biophysical signal models.
- The Modified Hodges detector is the most consistent detector across the different signal models and SNRs. It has an 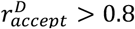 for the three signal models at 0dB and the biophysical model at -3dB SNR.
- The Fuzzy Entropy detector performs as well as the Modified Hodges detector for the Gaussian and biophysical signal models, but not on the Laplacian model.
- In terms of the average value of 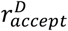, across the three signal models (last two columns of Table 3), the Modified Hodges detector performs the best for the 0dB signals, followed by the AGLR-G, AGLR-L, and Fuzzy Entropy detectors which have slightly lower but similar performance. For -3dB signals, the Modified Hodges, AGLR-G, and AGLR-L detectors result in similar performances.

Based on these observations, the Modified Hodges appears to be the most consistent detector for low SNR signal models, irrespective of the EMG signal model. The two statistical detectors (AGLR-G, and AGLR-L) and the Fuzzy Entropy detectors provide similar but slightly lower performance than the Modified Hodges detector.

## Discussion

Movement intention-triggered robot-assisted therapy is one of the options available for severely impaired patients without residual movement. EMG for movement intent detection is a simpler, more direct and task-specific alternative to EEG-BCI^23^. The investigation of EMG-triggered robot-assisted therapy requires a sensitive and robust method for the accurate and fast detection of movement intention from residual low SNR EMG signals. This study systematically investigated existing EMG detection algorithms in the literature until 2018. The investigation was carried out on simulated EMG signals using three different signal models with low SNR of 0dB and -3dB. These SNRs correspond to feeble EMG signals compared to regular surface EMG recordings from healthy individuals. Using three different signal models – two phenomenological and one biophysical – makes the study results robust to assumptions about the simulated EMG data.

The study by Staude et al. published in 2001^24^ compared different EMG detectors for accurate EMG onset-time detection. They employed a Gaussian signal model with ramp variance profiles (with varying slopes) at SNR of 3dB to 12dB in their analysis ^24^ and found the AGLR statistical detector to be the best in terms of onset detection, while the Hodges detector performed poorly^24^. Although there are some similarities between the current study and those of Staude et al., the two differ in several ways: (a) the current study is focused on realtime detection, while Staude et al.’s primary goal was offline analysis; (b) the current study employed lower SNR signals, which is important considering its application to detect motion intention in severally affected stroke patients; (c) the current study tested three different signal models, while Staude et al. used only the Gaussian signal model; (d) the primary performance measure in Staude et al. was onset detection latency, while the current study used a composite performance measure (or cost) consisting of the false positive rate, false negative rate, and detection latency; (e) the rationale for the choice of the specific detector parameters in Staude et al. was unclear. In the current study, the detector parameters were optimized through a brute force search to ensure the best detectors from each detector type were compared; and (f) the current study investigates a wider class of detector types than Staude et al., including the detectors published after 2001.

The current study identified that the Modified Hodges detector performed consistently well with cost *C* ≤ 0.2 for at least 80% of the validation trials, across the different signal models and SNRs, except for the -3dB Laplacian signal model, where all detectors fail. The Modified Hodges detector – a simplified version of the Hodges detector – performs better than Hodges because it does not involve the additional averaging step in computing its test function. This reduces the detection latency for the Modified Hodges detector without an appreciable increase in the false positive and false negative rates (Table S4 in the supplementary material). The AGLR-G, AGLR-L, and Fuzzy Entropy detectors perform slightly lower than Modified Hodges but better than the rest of the detectors. The good performance of the statistical detectors agrees with that of Staude et. al. even with the lower SNRs investigated in this study. The Fuzzy Entropy detector also performs well, unlike its counterpart – Sample Entropy. The Sample Entropy algorithm in this study used the local estimate of the signal’s standard deviation for normalizing the data. Sample entropy’s poor performance with the local estimate of the standard deviation was previously reported by Zhang et. al. Sample entropy performs well only with the global estimate of the signal’s standard deviation^31^. This is not suitable for real-time implementation as estimating the global standard deviation is a non-causal operation requiring the entire signal record. The use of the fuzzy similarity measure addresses this problem with Sample Entropy, allowing the Fuzzy Entropy detector to track changes in the overall signal amplitude. Interestingly, Fuzzy Entropy has a low cost of detection for both 0dB and -3dB biophysical signal models, which could be due to the additional structure of the MUAPs in the move-phase of the biophysical signal.

Interestingly, the RMS detector we used previously to demonstrate the viability of EMG as an alternative to detect movement intention in severely impaired chronic stroke subjects^23^ was not one of the best performers, as seen in Fig.5 and Table 3. We note that the observed performance was for the RMS detector with optimized parameters (Table 2) using the training dataset. This optimized RMS detector had a relatively high false negative rate and higher detection latency which resulted in its poor performance. This could possibly explain the lack of agreement between the EMG and EEG detectors we had observed in our previous study, and a more sensitive detector might have identified EMG activity in a larger proportion of subjects. The current study results warrant further investigation with real EMG data from severely impaired patients using other detectors, such as the Modified Hodges, AGLR-G/L, and Fuzzy Entropy.

In general, most detectors have a relatively lower cost of detection for the biophysical signal model, compared to the Gaussian and the Laplacian signal models. The reasons for the better performance on the biophysical model are not entirely clear, except for the Fuzzy Entropy detector. One possibility is the difference in the spectra of the signals from the biophysical model compared to the Gaussian or Laplacian modes (Fig. 1); more signal energy is concentrated in the lower frequencies for the biophysical model than in the Gaussian or Laplacian models. Most detectors compute their test functions through a lowpass filtering or averaging operation, which could be retaining a relatively larger portion of the signal in the biophysical model compared to the Gaussian and Laplacian ones, thus resulting in improved performance with the biophysical model. If this is correct, then the difference in performances between the biophysical and the Gaussian/Laplacian models should disappear when an appropriate spectral shaping filter is used in the Gaussian and Laplacian models, yielding a spectrum like the biophysical model. Finally, among the Gaussian and Laplacian models, the relatively poorer performance with the Laplacian signal model could be due to the long tails of the Laplacian distribution.

The simulated data used in the current study relies on a step-change in the signal properties between the rest- and move-phase, and an EMG signal of fixed amplitude during the move phase. These assumptions will be violated when dealing with feeble surface EMG signals recorded from impaired participants with no residual movements. In such participants, movement attempts are likely to produce intermittent bursts of EMG activity with smooth transitions between the on and off states in the target muscles. The EMG signal might have time-varying amplitude even when the participant can continuously activate the muscle for sufficient duration. Although based on idealized simulated EMG data, the current results do provide some idea about the detector types that can potentially work on real low SNR EMG signals; a detector performing poorly on ideal data is likely to perform worse with real data. Furthermore, the results from the current analysis also indicate that Modified Hodges, AGLR-G, AGLR-L, and Fuzzy Entropy detectors are likely to pick up even bursts of EMG signals since they have small detection latency (Δ*t* ≤ 50*ms*).

The detectors studied in this paper can only be used for on-off control of robotic assistance^47^, where once EMG activity is detected, robotic assistance drives the limb towards the target in a preprogrammed fashion. This is different from continuous control of the motion where robotic assistance is proportional to the level of EMG measured from the target muscles ^35^. Continuous control of robotic assistance is likely to be a more natural and engaging interaction for participants than simple on-off control. The most common EMG feature for continuous control has been the EMG amplitude estimate ^35^, however, the test functions employed by the different detector types in the current study could be potential alternatives. The choice of the best control variable depends on which one of these is sensitive, robust, and provides a natural human-robot interaction with minimal lag. However, it should be noted that it is unclear how well severely impaired participants, with no residual movements, can finely modulate their EMG activity. The choice of on-off versus continuous control of robotic assistance for a participant will require a screening procedure to evaluate the ability of the participant to modulate EMG activity in the target muscle.

The study has limitations that are worth noting to ensure that the results are interpreted appropriately. The study entirely relies on simulated data to investigate the different detectors. The conclusions are thus only as good as the assumed signal models and how well they represent the residual surface EMG signals of patients with no visible movements. This is the first study investigating detectors for low SNR EMG, and thus the use of simulated data was essential to gain some understanding of the performance of the different detectors. Simulated data also allows complete control of the ground truth, which provides a more truthful characterization of different detectors’ detection accuracy and latency. The use of three different signal models to investigate the different detectors also adds some robustness to the study’s findings. Additionally, this analysis allows us to exclude the poorly performing detectors and identify the ones that warrant further investigation with real data. Another potential limitation of the use of simulated data is the availability of complete information about the ground truth against which the different detectors are compared. However, the results of the current study can’t be verified with real data because we will never know the ground truth in the surface EMG from patients with no residual movements. This is a valid concern. Nevertheless, some form of an unsupervised approach will be required for verifying the results of the current study with real data. With real data, the best detector would be the one that consistently provides the maximum separation for the probability density function of the test function *g*[*n*] from the different detectors under the rest-phase and move-phase.

## Conclusion

This paper systematically investigated existing EMG detection algorithms on low SNR EMG signals simulated using three different signal models (two phenomenological – Gaussian, Laplacian models and a biophysical model) at two different SNRs (0dB and -3dB). The Modified Hodges detector – a simplified version of the threshold-based Hodges detector, introduced in the current study – was found to be the most consistent detector across the different signal models and SNRs. This detector had false positive and false negative rates of lower than 20% and a detection latency of lower than 50ms for almost 90% of the trials on which it was tested for 0 dB SNR and more than 40% of the trials for -3 dB SNR. The two statistical detectors (Gaussian and Laplacian Approximate Generalized Likelihood Ratio) and the Fuzzy Entropy detectors have a slightly lower performance than Modified Hodges. Overall, the Modified Hodges, Gaussian and Laplacian Approximate Generalized Likelihood ratio, and Fuzzy entropy detectors were identified as potential candidates for further validation with real surface EMG data on a population of severely impaired patients. The current study forms the first step towards developing a simpler, practical, and robust EMG-based human-machine interface for triggered robot-assisted therapy in severely impaired patients.

## Supporting information

Supplemental Table S1, S2, S3 and S4 will be used for the link to the file on the preprint site.

## Data Availability

All data produced are available online at https://github.com/1608Moni/EMG_detectors

## Acknowledgements

The authors thank Prof. Suresh Devasahayam for providing us with the software he’d developed for simulating the EMG signals. This software was used for generating data for the biophysical EMG model.

## Author contributions

SB conceived and brainstormed the idea with EB and VSKM. PR worked on the initial literature search and the implementation of some of the detectors. MY implemented the different EMG signal models, all the detectors, optimization of detector parameters, and the analysis of the results. AD and SB provided scientific inputs for the implementation and analysis done by MY. EB and VSKM provided critical feedback for the methodology and results. MY, AD, and SB wrote the first draft of the manuscript. All authors reviewed and approved the final submitted manuscript.

## Competing interest statement

The authors declare that they do not have any competing interests.

### Algorithm 1

Procedure for selecting the best parameter combination for the detector type.

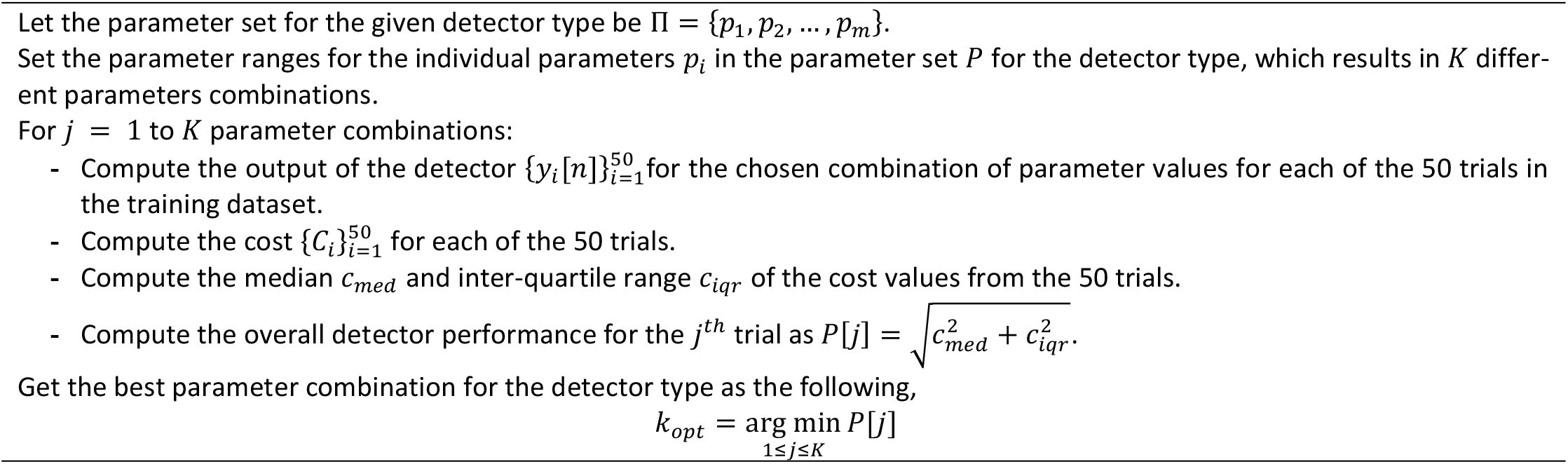

