## Supplemental Table S1, S2, S3 and S4 will be used for the link to the file on the preprint site. for "A Systematic Investigation of Detectors for Low Signal-to-Noise Ratio EMG Signals"

**Supplementary Material**

| **Table S1:** Set of parameter ranges used for optimizing the detector parameters using the training dataset. | | | |
| --- | --- | --- | --- |
|  | **Parameters** | **Range** | **Step size** |
| **Hodges** | Window size (ms) | $\left[ 100 500 \right]$ | 50 |
|  | Weights for threshold | $\left[ 1 5 \right]$ | 1 |
|  | LPF Cut-off frequency (Hz) | $\left[ 0.5 10 \right]$ | 1 |
| **Modified Hodges** | Weights for threshold | $\left[ 1 5 \right]$ | 1 |
|  | LPF Cut-off frequency (Hz) | $\left[ 0.5 10 \right]$ | 1 |
| **Modified Lidierth** | Weights for threshold | $\left[ 1 5 \right]$ | 1 |
|  | LPF Cut-off frequency (Hz) | $\left[ 0.5 10 \right]$ | 1 |
|  | T1 (ms) | (30,60,100) | - |
|  | m (ms) | [5 100] : m < T1 | 5 |
| **Lidierth** | Window size (ms) | $\left[ 100 500 \right]$ | 50 |
|  | Weights for threshold | $\left[ 1 5 \right]$ | 1 |
|  | T1 (ms) | (30,60,100) | - |
|  | m (ms) | [5 100] : m < T1 | 5 |
| **Bonato** | Weights for threshold | $\left[ 1 5 \right]$ | 1 |
|  | T1 (ms) | (30,60,100) | - |
|  | m (ms) | [5 100] : m < T1 | 5 |
| **TKEO** | Window size (ms) | $\left[ 100 500 \right]$ | 50 |
|  | Weights for threshold | $\left[ 1 5 \right]$ | 1 |
|  | HPF Cut-off frequency (Hz) | $\left[ 5 30 \right]$ | 5 |
|  | T1 (ms) | (30,60,100) | - |
| **AGLR-G** | Window size (ms) | $\left[ 100 500 \right]$ | 50 |
|  | Weights for threshold | $\left[ 1 5 \right]$ | 1 |
| **AGLR-L** | Window size (ms) | $\left[ 100 500 \right]$ | 50 |
|  | Weights for threshold | $\left[ 1 5 \right]$ | 1 |
| **Fuzzy Entropy** | Window size (ms) | $\left[ 100 500 \right]$ | 50 |
|  | Threshold | $\left[ 1 5 \right]$ | 1 |
| **Sample Entropy** | Window size (ms) | [50 150] | 50 |
|  | Threshold | $\left[ 1 5 \right]$ | 1 |
|  | Tolerance for distance | [0.5 2.5] | 1 |
| **CWT** | Weights | [1 2] | 0.1 |
| **SSA** | Window size (ms) | [50 100] | 2 |
| **RMS** | Window Size (ms) | $\left[ 120 440 \right]$ | 40 |
|  | Weight | $\left[ 1 5 \right]$ | 1 |
|  | Window shift (ms) | (1, 10, 20,40) | - |
|  | Time threshold (ms) | $\left[ 40 200 \right]$ | 40 |

**Algorithms for the different detectors investigated in the study**

**Detector 1: Hodges detector**

Parameters

- Weights	$\alpha$
- LPF cutoff frequency $\mathrm{Fc}\left( \mathrm{Hz} \right)$
- Window size (ms) $W$

$$x\left[ n \right] =Recorded EMG signal$$

- $\tilde{x}\left[ n \right]=filter\left( \left| x\left[ n \right] \right| \right) ;filter\left( \cdot\right) i$s the filter function (2^nd^ order Butter worth causal low pass filter)
- Compute the mean and standard deviation of the baseline: $\left( \mu_{g},\sigma_{g} \right)=T(\left\{ \tilde{x}\left[ n \right] \right\}_{k=1}^{M})$
- Compute the threshold using the baseline parameters: $h=\mu_{g}+\alpha\cdot\sigma_{g},$

For n $\geq$ Baseline window

- Compute the moving average in sliding window $\tilde{y}\left[ n \right]= \frac{1}{W}\sum_{k=n-W+1}^{n} \tilde{x}\left[ n \right]$
- Compute the test function: $g\left[ n \right]= \frac{1}{\sigma_{g}}\left( \tilde{y}\left[ n \right]-\mu_{g} \right)$
- Obtain the binary output and estimate the onset time of the detector as follows:
- $y\left[ n \right]=\left\{ \begin{matrix} 1 & g\left[ n \right]>\alpha\\ 0 & g\left[ n \right]\leq\alpha\end{matrix} \right.$
  - $\hat{t_{0}}=min\left\{ n | y\left[ n \right]=1; N_{r}\leq n<N_{t} \right\}$

**Detector 2: Modified Hodges detector**

Parameters

- Weights	$\alpha$
- LPF cutoff frequency $\mathrm{Fc}\left( \mathrm{Hz} \right)$

$x\left[ n \right] =Recorded EMG signal$ :

- $\tilde{x}\left[ n \right]=filter\left( \left| x\left[ n \right] \right| \right) ;filter\left( \cdot\right) i$s the filter function (2^nd^ order Butter worth causal low pass filter)
- Compute the mean and standard deviation of the baseline: $\left( \mu_{g},\sigma_{g} \right)=T(\left\{ \tilde{x}\left[ n \right] \right\}_{k=1}^{M})$
- Compute the threshold using the baseline parameters: $h=\mu_{g}+\alpha\cdot\sigma_{g},$

For n $\geq$ Baseline window

- Compute the test function: $g\left[ n \right]= \tilde{x}\left[ n \right]$
- Obtain the binary output and estimate the onset time of the detector as follows:
- $y\left[ n \right]=\left\{ \begin{matrix} 1 & g\left[ n \right]>h \\ 0 & g\left[ n \right]\leq h \end{matrix} \right.$
- $\hat{t_{0}}=min\left\{ n | y\left[ n \right]=1; N_{r}\leq n<N_{t} \right\}$

**Detector 3: Modified Lidierth**

Parameters:

- Weights	$\alpha$
- LPF cutoff frequency $\mathrm{Fc}\left( \mathrm{Hz} \right)$
- $T_{1}$ (ms),
- $m$ (ms)

$x\left[ n \right] =Recorded EMG signal$ :

Compute the test function as in Modified hodges.

Obtain the binary output and estimate the onset time of the detector as follows:

First thresholding

$$b1\left[ n \right]=\left\{ \begin{matrix} 1 & g\left[ n \right]>h \\ 0 & g\left[ n \right]\leq h \end{matrix} \right.$$

- $y\left[ n \right]=DoubleThreshold(b1\left[ n \right],m,T_{1})$
- $\hat{t_{0}}=min\left\{ n | y\left[ n \right]=1; N_{r}\leq n<N_{t} \right\}$

**Detector 4: Lidierth**

Parameters:

- Weights $\alpha$
- Window size (ms) $W$
- $T_{1}$ (ms)
- $m$ (ms)

$x\left[ n \right] =Recorded EMG signal$:

Compute the test function as in hodges.

Obtain the binary output and estimate the onset time of the detector as follows:

- $b1\left[ n \right]=\left\{ \begin{matrix} 1 & g\left[ n \right]>h \\ 0 & g\left[ n \right]\leq h \end{matrix} \right.$
- $y\left[ n \right]=DoubleThreshold(b1\left[ n \right],m,T_{1})$
- $\hat{t_{0}}=min\left\{ n | y\left[ n \right]=1; N_{r}\leq n<N_{t} \right\}$

**Detector 5: Bonato**

Parameters:

- Weights $\alpha$
- Window size (ms) $W$
- Baseline window (ms)	M

$$x\left[ n \right]=Recorded EMG signal$$

$\tilde{x}\left[ n \right]=filter\left( x\left[ n \right] \right) ; filter\left( \cdot\right)-8th order Adaptive whitening filter$

For $n=1 to N_{t}$

- $g\left[ n \right]=\left( \tilde{x}^{2}\left[ n \right]-\tilde{x}^{2}\left[ n-1 \right] \right)/{\sigma_{0}^{2}};n\in\left\{ 1, 3, 5, \ldots\right\}$If $n>M$
- Compute the mean and standard deviation of the baseline: $\left( \mu_{g},\sigma_{g} \right)=T(\left\{ g\left[ n \right] \right\}_{k=1}^{M})$
- Compute the threshold using the baseline parameters: $h=\mu_{g}+\alpha\cdot\sigma_{g},$

$b1\left[ n \right]=\left\{ \begin{matrix} 1 & g\left[ n \right]>h \\ 0 & g\left[ n \right]\leq h \end{matrix} \right.$

- $y\left[ n \right]=DoubleThreshold(b1\left[ n \right],1,T_{1})$
- $\hat{t_{0}}=min\left\{ n | y\left[ n \right]=1; N_{r}\leq n<N_{t} \right\}$

**Detector 6: TKEO**

Parameters:

- Weights $\alpha$
- Window size (ms) $W$
- $T_{1}$ (ms)
- $High pass filter cut-off freq$
- Baseline window(ms)	M

$$x\left[ n \right]=Recorded EMG signal$$

$\tilde{x}\left[ n \right]=filter\left( x\left[ n \right] \right) ; filter\left( \cdot\right)-6th order High pass filter$

For $n=1 to N_{t}$

- $\varphi[n]=\tilde{x}^{2}\left[ n-1 \right]-\tilde{x}^{2}\left[ n-2 \right]\tilde{x}^{2}\left[ n \right]$
- $g\left[ n \right]=\frac{1}{W}\sum_{k=n-W+1}^{n} \varphi\left[ k \right]$
- If $n>M$
- Compute the mean and standard deviation of the baseline: $\left( \mu_{g},\sigma_{g} \right)=T(\left\{ g\left[ n \right] \right\}_{k=1}^{M})$
- Compute the threshold using the baseline parameters: $h=\mu_{g}+\alpha\cdot\sigma_{g},$

$b1\left[ n \right]=\left\{ \begin{matrix} 1 & g\left[ n \right]>h \\ 0 & g\left[ n \right]\leq h \end{matrix} \right.$

- $y\left[ n \right]=DoubleThreshold(b1\left[ n \right],1,T_{1})$
- $\hat{t_{0}}=min\left\{ n | y\left[ n \right]=1; N_{r}\leq n<N_{t} \right\}$

**Detector 7: Approximate generalized likelihood ratio test (AGLR-G)**

Parameters:

- Weights $\alpha$
- Window size (ms) $W$
- Baseline window (ms)	M

$$x\left[ n \right]=Recorded EMG signal$$

$\tilde{x}\left[ n \right]=filter\left( x\left[ n \right] \right) ; filter\left( \cdot\right)-8th order Adaptive whitening filter$

Maximum likelihood estimates of the variance are derived assuming the step variance of gaussian model.

$\hat{\theta_{0}}= \frac{1}{M}\sum_{i=1}^{M} \tilde{x}^{2}\left[ i \right]$

For $n=1 to N_{t}$

- $\hat{\theta_{1}}\left( n \right)= \frac{1}{W}\sum_{k=n-W+1}^{n} \tilde{x}^{2}\left[ k \right]$
- $\hat{\rho}\left( n \right)= \frac{\hat{\theta_{1}}\left( n \right)}{\hat{\theta_{0}}}$
- $g\left[ n \right]=\frac{W}{2}\left( \hat{\rho}\left( n \right)-\ln\hat{\rho}\left( n \right)-1 \right)$
- If $n>Baseline window$
- Compute the mean and standard deviation of the baseline: $\left( \mu_{g},\sigma_{g} \right)=T(\left\{ g\left[ n \right] \right\}_{k=1}^{M})$
- Compute the threshold using the baseline parameters: $h=\mu_{g}+\alpha\cdot\sigma_{g},$

$y\left[ n \right]=\left\{ \begin{matrix} 1 & g\left[ n \right]>h \\ 0 & g\left[ n \right]\leq h \end{matrix} \right.$

- $\hat{t_{0}}=min\left\{ n | y\left[ n \right]=1; N_{r}\leq n<N_{t} \right\}$

**Detector 8: Approximate generalized likelihood ratio test Laplace (AGLR- L)**

Parameters:

- Weights $\alpha$
- Window size (ms) $W$

$$x\left[ n \right]=Recorded EMG signal$$

$\tilde{x}\left[ n \right]=filter\left( x\left[ n \right] \right) ; filter\left( \cdot\right)-8th order Adaptive whitening filter$

Maximum likelihood estimates of the variance are derived assuming the step variance of laplacian model.

$\hat{\theta_{0}}= \frac{\sqrt{2}}{M}\sum_{i=1}^{M} \left| \tilde{x}\left[ i \right] \right|$

For $n=1 to N_{t}$

- $\hat{\theta_{1}}\left( n \right)= \frac{\sqrt{2}}{W}\sum_{k=n-W+1}^{n} \left| \tilde{x}\left[ k \right] \right|$
- $\hat{\rho}\left( n \right)= \frac{\hat{\theta_{1}}\left( n \right)}{\hat{\theta_{0}}}$
- $g\left[ n \right]=\frac{W}{2}\left( \hat{\rho}\left( n \right)-\ln\hat{\rho}\left( n \right)-1 \right)$
- If $n>M$
- Compute the mean and standard deviation of the baseline: $\left( \mu_{g},\sigma_{g} \right)=T(\left\{ g\left[ n \right] \right\}_{k=1}^{M})$
- Compute the threshold using the baseline parameters: $h=\mu_{g}+\alpha\cdot\sigma_{g},$

$y\left[ n \right]=\left\{ \begin{matrix} 1 & g\left[ n \right]>h \\ 0 & g\left[ n \right]\leq h \end{matrix} \right.$

- $\hat{t_{0}}=min\left\{ n | y\left[ n \right]=1; N_{r}\leq n<N_{t} \right\}$

**Detector 9: Sample Entropy**

Parameters:

- Tolerance	$\rho$
- Threshold $h$
- Window size (ms) $W$
- Embedding dimension (m) = 2

$$x\left[ n \right]=Recorded EMG signal$$

- If n > W
- $Seg[n]=\left\{ x\left[ n \right] \right\}_{k=n-W+1}^{n}$
- r = $\rho*stdv(Seg[n])$
- $X_{m}\left( p \right)=\left[ Seg\left( p+k \right) \right]_{k=0}^{m-1} p=1,2,3\ldots.W-m$
- Calculate the Chebyshev distance excluding the self-match case.
- ${d\left[ X_{m}\left( p \right),X_{m}\left( q \right) \right]}_{p\neq q}= \max_{q} \left| X\left( p \right)-X\left( q \right) \right|$
- $B_{m}\left( r \right)$= probability that two sequences match for ‘m’ points: $d\left[ X_{m}\left( p \right)X_{m}\left( q \right) \right]<r$
- $A_{m}\left( r \right)$ = probability calculated for ‘m+1’ dimension. : $d\left[ X_{m+1}\left( p \right)X_{m+1}\left( q \right) \right]<r$
- $g\left[ n \right]= -\ln\left( {A_{m}\left( r \right)}/{B_{m}\left( r \right)} \right)$
- If $n>Baseline\_window$
- Compute the mean and standard deviation of the baseline: $\left( \mu_{g},\sigma_{g} \right)=T(\left\{ g\left[ n \right] \right\}_{k=1}^{M})$
- Compute the threshold using the baseline parameters: $h=\mu_{g}+\alpha\cdot\sigma_{g},$

$y\left[ n \right]=\left\{ \begin{matrix} 1 & g\left[ n \right]>h \\ 0 & g\left[ n \right]\leq h \end{matrix} \right.$

- $\hat{t_{0}}=min\left\{ n | y\left[ n \right]=1; N_{r}\leq n<N_{t} \right\}$

**Detector 10: Fuzzy Entropy**

Parameters:

- Embedding dimension (m)	= 2
- Tolerance (r)	= 0.25
- Fuzzy function parameter (l) = 2
- Threshold $h$
- Window size (ms) $W$

$$x\left[ n \right]=Recorded EMG signal$$

- If n > W
- $Seg[n]=\left\{ x\left[ n \right] \right\}_{k=n-W+1}^{n}$
- $X_{m}\left( i \right)=\left[ Seg\left( i+k \right) \right]_{k=0}^{m-1} i=1,2,3\ldots.W-m$
- Calculate the Chebyshev distance excluding the self-match case.
- ${d_{ij}=d\left[ X_{m}\left( i \right),X_{m}\left( j \right) \right]}_{i\neq j}= \max_{j} \left| X\left( i \right)-X\left( j \right) \right|$
- $D_{ij}^{m}= exp(-{\left( d_{ij}^{m} \right)^{n}}/r)$
- Compute the probability that the vector $X_{j}^{m}$ is within the similarity tolerance r of template vector $X_{i}^{m}$:
- $C_{m}\left( l,r \right)= \frac{1}{W-m}\sum_{i=1}^{W-m} \left( \frac{1}{W-m-1}\sum_{j=1,j\neq i}^{N-m} D_{ij}^{m} \right)$
- $g\left[ n \right]= \ln\left( {C_{m}\left( l,r \right)}/{C_{m+1}\left( l,r \right)} \right)$
- If $n>M$
- Compute the mean and standard deviation of the baseline: $\left( \mu_{g},\sigma_{g} \right)=T(\left\{ g\left[ n \right] \right\}_{k=1}^{M})$
- Compute the threshold using the baseline parameters:$h=\mu_{g}+\alpha\cdot\sigma_{g},$

$y\left[ n \right]=\left\{ \begin{matrix} 1 & g\left[ n \right]>h \\ 0 & g\left[ n \right]\leq h \end{matrix} \right.$

- $\hat{t_{0}}=min\left\{ n | y\left[ n \right]=1; N_{r}\leq n<N_{t} \right\}$

**Detector 11: Singular spectrum analysis**

Parameters:

- Window size	m
- Lag parameter: M = m/2
- Test interval parameter: p = m-M+1
- Test interval parameter: q = m+1

$$x\left[ n \right]=Recorded EMG signal$$

Segment the signal of window size m and form a trajectory matrix $X_{n}$ with the lag parameter M.

- Compute the lag covariance matrix ${R=X}_{n}*X_{n}^{T}$
- Find the eigenvalues and eigenvectors for $R .$
- L = no. of eigen vectors with eigen value > 5% of $\sum eigenvalue of R$.
- Arrange the eigenvectors in descending order and choose the first L eigen vectors and form the matrix $U$.
- Form a test matrix $T_{n}$with new set of points.
- Test function: $D_{n}=\sum_{j=p+1}^{q} \left( \left( T_{j}^{\left( n \right)} \right)^{T}T_{j}^{\left( n \right)}-\left( T_{j}^{\left( n \right)} \right)^{T}UU^{T}T_{j}^{\left( n \right)} \right)$ , Where $T_{j}^{\left( n \right)}$ column vectors of test matrix $T_{n}$.
- Compute ${(C}_{n})$CUSUM statistics (ref [xxx]).
- $\hat{t_{0}}$= First point with zero value before CUSUM statistics reaches maximum.
- $Compute peaks in the$ ${(C}_{n})$ $N_{r}$ <n< $N_{t}$.
- $y\left[ n \right]=1$. If n+1 is peak of ${(C}_{n}).$First location where the statistics is equal to zero preceding the peak time.

**Detector 12: Continuous wavelet transform**

Parameters:

- MUAP constant $k_{n,1}$ =1
- MUAP wavelet duration ( $\tau$) = 200 ms
- MUAP wavelet parameter ($\lambda$) = [0.5, 1.5, 2, 3]
- Scaling factor for wavelet (a) = [1,3,4,6]
- Wavelet function: $wav\left[ n \right]=k_{n,1}.2\left( \frac{n}{\lambda_{n}} \right)e^{-\frac{k^{2}}{\lambda^{2}}}$
- Weights	α

$$x\left[ n \right]=Recorded EMG signal$$

- If $n>Baseline\_window$
- $CWT (a,\tau)$ = $\sum_{-\infty}^{+\infty} x\left[ n \right]. {wav}^{*}\left[ \frac{n-\tau}{a} \right]$
- $g\left[ n \right]=\min_{a} CWT (a,\tau)$
- $\zeta=max\left\{ g\left[ n \right] \right\}$
- $h=\alpha.\zeta$

$y\left[ n \right]=\left\{ \begin{matrix} 1 & g\left[ n \right]>h \\ 0 & g\left[ n \right]\leq h \end{matrix} \right.$

- $\hat{t_{0}}=min\left\{ n | y\left[ n \right]=1; N_{r}\leq n<N_{t} \right\}$

**Detector 13: Root Mean Square Detector**

Parameters:

- Weights	α
- Window size(ms)	W
- Window shift(ms)	p
- Temporal Threshold(ms)	T1
- Temp Threshold (samples)	m = T1/p

$$x\left[ n \right]=Recorded EMG signal$$

- $\tilde{x}\left[ n \right]=filter\left( \left| x\left[ n \right] \right| \right) ;filter\left( \cdot\right) i$s the filter function (2^nd^ order Butter worth causal band pass filter)
- If $n>Window size$
- $g\left[ n \right]= \sqrt{\frac{1}{W}\sum_{k=n-W+1}^{n} \tilde{x}^{2}\left[ k \right]}$
- $n=n+p$
- b1$\left[ n \right]=\left\{ \begin{matrix} 1 & g\left[ n \right]>h \\ 0 & g\left[ n \right]\leq h \end{matrix} \right.$
- $y\left[ 0 \right]=0$
- F1[n] = $\frac{1}{m}\sum_{i=n-m+1}^{m} b1\left[ i \right]$
- y[n]= $\left\{ \begin{aligned} 1 if F1\left[ n \right]=1 \\ 0 if F1\left[ n \right]=0 \\ y\left[ n-1 \right] if 0<F1\left[ n \right]<1 \end{aligned} \right.$
- $\hat{t_{0}}=min\left\{ n | y\left[ n \right]=1; N_{r}\leq n<N_{t} \right\}$

**Algorithm S1: Double Threshold Algorithm**

Input: Binary signal after first thresholding (b1), Double threshold parameters (m, $T_{1}$)

Output: Binary output of the detector $y\left[ n \right]$

- To verify at least n out of m samples crosses the threshold – “Active state.”
- For n > m

F1[n] = $\frac{1}{m}\sum_{i=n-m+1}^{m} b1\left[ i \right]$

$b2\left[ n \right]=\left\{ \begin{matrix} 1 & F1\left[ n \right]>0 \\ 0 & F1\left[ n \right]\leq0 \end{matrix} \right.$

- To verify that such an Active state lasts for $T_{1}$ samples.
- For n > $T_{1}$

F2[n] = $\frac{1}{T_{1}}\sum_{i=n-T_{1}+1}^{T_{1}} b2\left[ i \right]$

- $y\left[ n \right]=\left\{ \begin{matrix} 1 & F2\left[ n \right]=1 \\ 0 & Otherwise \end{matrix} \right.$

**Algorithm S2: Adaptive whitening algorithm**

Input: Raw EMG signal

Output: Adaptive whitened signal.

Order of filter (p) = 8

Auto regressive model driven by gaussian noise is defined as follows:

- $z[n] = \sum_{k=1}^{p} a\left[ k \right]z\left[ n-k \right]+ e[n]$
- Where e[n] whiten noise with zero mean and variance $\sigma^{2}$
- Arrange the time series into M linear equation
- Compute the least square estimate of the parameter vector a
- $\tilde{x}\left[ n \right]= x\left[ n \right]- \sum_{k=1}^{p} \hat{a}\left[ k \right]x\left[ n-k \right]$

**Table S2**: Two-way ANOVA for individual signal model testing the effect of algorithm and SNR on the cost.

Gaussian

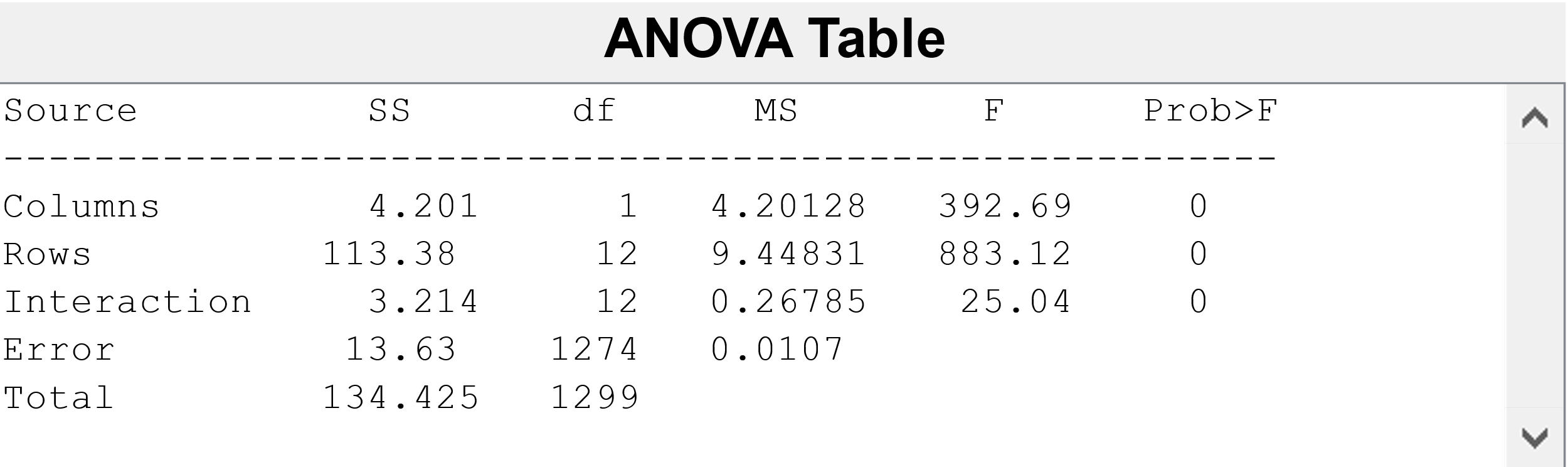

Laplacian

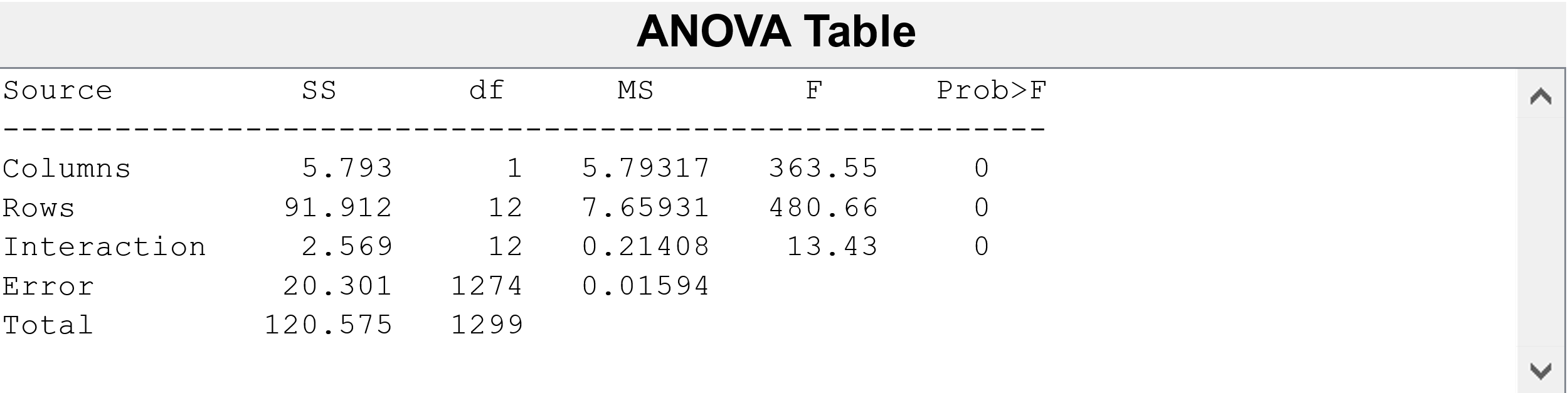

Biophysical

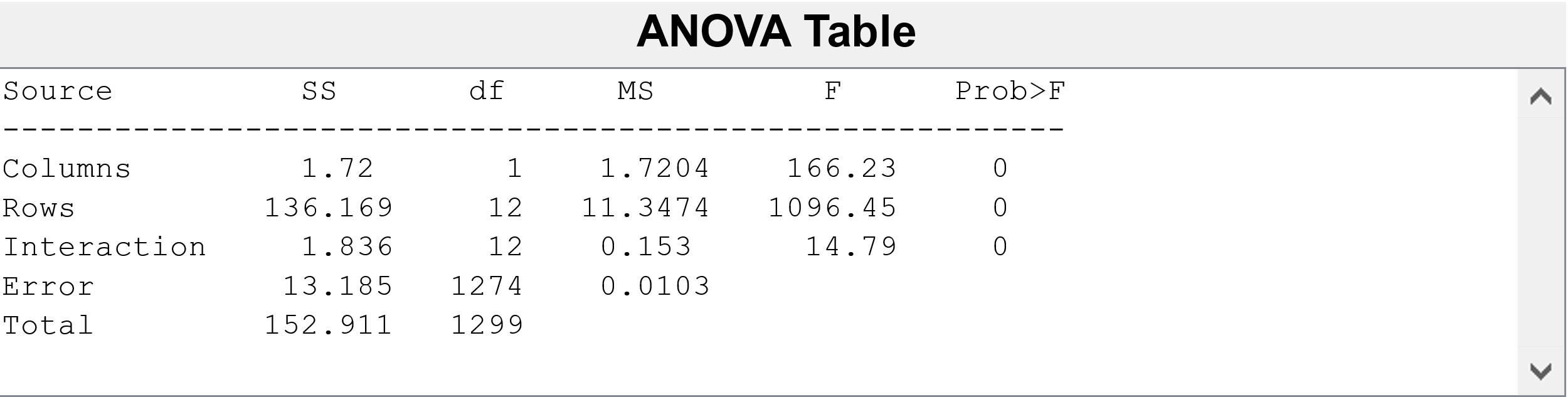

**Table S3**: Parameters used by the biophysical model-based simulator the generate the EMG data

- Maximum fibers per motor unit	320
- Maximum motor unit	32
- Fiber Action potential Width	64
- Surface electrode Width (mm)	25
- Inter-site separation (mm)	30
- Surface Muscle Depth_(mm)	30
- SurfElectDist0 (mm)	40

(Distance from muscle origin)

- Muscle Length (mm)	100
- Muscle Diameter (mm)	16
- Surface Electrode Diameter (mm)	4
- Gain Electrode	1

(Bipolar 2 channel Electrode configuration)

- Number of muscle fibers for each motor unit : [32 144] with step size of 4 each value repeated twise
- Conduction velocity: {4,4,4, 4, 5, 5, 5, 5, 6, 6, 6, 6, 7, 7, 7, 7, 8, 8, 8, 8, 8, 8, 8, 8, 10, 10, 10, 10, 10, 10, 10,10 }

| **Table S4 :** Mean and standard deviation of the false positive rate, false negative rate, Latency and cost of the Detectors | | | | | | | | |
| --- | --- | --- | --- | --- | --- | --- | --- | --- |
| **Gaussian** | | | | | | | | |
|  | **FNR** | | **FPR** | | **Latency (ms)** | | **Cost** | |
|  | **0 dB** | **-3 dB** | **0 dB** | **-3 dB** | **0 dB** | **-3 dB** | **0 dB** | **-3 dB** |
| **Modified Hodges** | 0.024±0.013 | 0.208±0.05 | 0.125±0.039 | 0.103±0.036 | 23.52±16.035 | 34.52±31.855 | 0.144±0.038 | 0.235±0.082 |
| **AGLR-G** | 0.009±0.007 | 0.096±0.041 | 0.07±0.035 | 0.147±0.058 | 26.1±23.044 | 32.62±32.04 | 0.123±0.081 | 0.206±0.084 |
| **AGLR-L** | 0.012±0.008 | 0.12±0.051 | 0.071±0.039 | 0.144±0.057 | 32.88±23.838 | 37.56±36.317 | 0.146±0.084 | 0.222±0.1 |
| **Fuzzy Entropy** | 0.119±0.029 | 0.376±0.07 | 0.089±0.029 | 0.039±0.024 | 27.46±17.882 | 58.64±45.604 | 0.149±0.042 | 0.401±0.103 |
| **Modified Lidierth** | 0.042±0.018 | 0.261±0.054 | 0.102±0.033 | 0.113±0.033 | 36.42±26.564 | 45.1±33.249 | 0.178±0.069 | 0.283±0.069 |
| **Hodges** | 0.013±0.007 | 0.18±0.058 | 0.052±0.035 | 0.04±0.026 | 55.56±27.458 | 76.2±45.563 | 0.227±0.102 | 0.328±0.154 |
| **RMS** | 0.063±0.028 | 0.466±0.102 | 0.014±0.013 | 0.011±0.012 | 68.8±37.939 | 204±173.934 | 0.322±0.186 | 0.686±0.246 |
| **Lidierth** | 0.028±0.016 | 0.265±0.069 | 0.016±0.016 | 0.021±0.018 | 68.9±25.377 | 92.98±60.764 | 0.276±0.1 | 0.402±0.18 |
| **TKEO** | 0.031±0.018 | 0.308±0.076 | 0.052±0.034 | 0.044±0.025 | 54.32±27.515 | 77.62±55.712 | 0.224±0.098 | 0.396±0.155 |
| **Bonato** | 0.122±0.038 | 0.273±0.06 | 0.151±0.038 | 0.186±0.046 | 34.88±30.698 | 47.68±38.338 | 0.203±0.07 | 0.316±0.069 |
| **Sample Entropy** | 0.882±0.029 | 0.88±0.03 | 0.122±0.025 | 0.122±0.031 | 57.14±52.527 | 44.7±50.176 | 0.883±0.029 | 0.882±0.034 |

| **Biophysical** | | | | | | | | |
| --- | --- | --- | --- | --- | --- | --- | --- | --- |
|  | **FNR** | | **FPR** | | **Latency (ms)** | | **Cost** | |
|  | **0 dB** | **-3 dB** | **0 dB** | **-3 dB** | **0 dB** | **-3 dB** | **0 dB** | **-3 dB** |
| **Modified Hodges** | 0.006±0.005 | 0.104±0.034 | 0.081±0.045 | 0.069±0.032 | 19.18±10.187 | 35.54±37.726 | 0.102±0.04 | 0.169±0.138 |
| **AGLR-G** | 0.037±0.03 | 0.118±0.055 | 0.045±0.042 | 0.145±0.051 | 34.5±35.314 | 31.92±42.044 | 0.155±0.131 | 0.216±0.129 |
| **AGLR-L** | 0.063±0.035 | 0.089±0.051 | 0.146±0.054 | 0.169±0.063 | 20.8±23.772 | 50.74±61.079 | 0.172±0.068 | 0.286±0.197 |
| **Fuzzy Entropy** | 0.033±0.022 | 0.082±0.033 | 0.007±0.007 | 0.099±0.046 | 15.48±15.248 | 11.3±14.106 | 0.07±0.057 | 0.125±0.046 |
| **Modified Lidierth** | 0.089±0.035 | 0.17±0.037 | 0.049±0.024 | 0.099±0.032 | 36.5±9.788 | 44.36±31.76 | 0.15±0.033 | 0.209±0.105 |
| **Hodges** | 0.009±0.005 | 0.116±0.04 | 0.053±0.035 | 0.041±0.025 | 38.18±17.269 | 57.3±42.021 | 0.155±0.065 | 0.234±0.163 |
| **RMS** | 0.009±0.003 | 0.067±0.029 | 0.01±0.013 | 0.012±0.016 | 44.8±12.493 | 53.6±30.889 | 0.179±0.05 | 0.238±0.156 |
| **Lidierth** | 0.027±0.018 | 0.161±0.047 | 0.011±0.011 | 0.026±0.019 | 48.32±19.473 | 65.22±43.283 | 0.193±0.078 | 0.266±0.169 |
| **TKEO** | 0.105±0.045 | 0.472±0.087 | 0.047±0.032 | 0.045±0.03 | 58.6±37.56 | 99.3±89.364 | 0.237±0.148 | 0.54±0.168 |
| **Bonato** | 0.215±0.061 | 0.224±0.059 | 0.185±0.054 | 0.326±0.067 | 43.76±33.228 | 31.48±32.355 | 0.266±0.076 | 0.342±0.064 |
| **Sample Entropy** | 0.967±0.012 | 0.94±0.019 | 0.121±0.035 | 0.112±0.027 | 176.46±239.641 | 106.98±102.771 | 0.972±0.018 | 0.944±0.025 |

| **Laplacian** | | | | | | | | |
| --- | --- | --- | --- | --- | --- | --- | --- | --- |
|  | **FNR** | | **FPR** | | **Latency (ms)** | | **Cost** | |
|  | **0 dB** | **-3 dB** | **0 dB** | **-3 dB** | **0 dB** | **-3 dB** | **0 dB** | **-3 dB** |
| **Modified Hodges** | 0.08±0.04 | 0.292±0.073 | 0.144±0.049 | 0.119±0.038 | 28.08±18.489 | 53.5±39.173 | 0.169±0.057 | 0.324±0.105 |
| **AGLR-G** | 0.063±0.039 | 0.232±0.092 | 0.138±0.045 | 0.172±0.082 | 30.5±29.676 | 58.1±58.867 | 0.183±0.086 | 0.325±0.162 |
| **AGLR-L** | 0.041±0.027 | 0.191±0.082 | 0.144±0.049 | 0.168±0.091 | 28.52±29.017 | 55.78±52.62 | 0.186±0.078 | 0.308±0.161 |
| **Fuzzy Entropy** | 0.15±0.06 | 0.485±0.076 | 0.1±0.042 | 0.112±0.031 | 41.96±29.337 | 45.3±41.672 | 0.214±0.086 | 0.491±0.095 |
| **Modified Lidierth** | 0.1±0.047 | 0.262±0.074 | 0.137±0.043 | 0.158±0.045 | 44.94±26.25 | 66.5±44.907 | 0.214±0.065 | 0.345±0.103 |
| **Hodges** | 0.042±0.031 | 0.306±0.087 | 0.063±0.042 | 0.064±0.032 | 67.12±27.721 | 103.4±66.88 | 0.273±0.102 | 0.451±0.212 |
| **RMS** | 0.107±0.066 | 0.492±0.112 | 0.044±0.038 | 0.047±0.03 | 95.2±59.048 | 176.8±224.694 | 0.397±0.221 | 0.641±0.228 |
| **Lidierth** | 0.071±0.047 | 0.318±0.084 | 0.036±0.028 | 0.066±0.032 | 73.1±28.541 | 97.86±60.182 | 0.296±0.107 | 0.439±0.187 |
| **TKEO** | 0.129±0.066 | 0.463±0.089 | 0.066±0.039 | 0.065±0.033 | 72.14±41.693 | 108.08±75.708 | 0.3±0.151 | 0.542±0.168 |
| **Bonato** | 0.146±0.068 | 0.268±0.086 | 0.261±0.081 | 0.306±0.089 | 48.64±52.822 | 67.2±82.626 | 0.322±0.13 | 0.431±0.171 |
| **Sample Entropy** | 0.873±0.035 | 0.873±0.031 | 0.123±0.04 | 0.132±0.029 | 75±80.009 | 58.38±72.279 | 0.879±0.045 | 0.878±0.04 |
